# Effect of Neck-Bending on Upper Airway Caliber and Surrounding Soft Tissues in Controls and Apneics

**DOI:** 10.1101/2025.06.05.25329072

**Authors:** Richard J Schwab, Theodore C Lin, Andrew S Wiemken, Si Hao Tang, Allison Schwab, Brendan T Keenan

**Affiliations:** Division of Sleep Medicine, Department of Medicine, University of Pennsylvania, Philadelphia, PA

**Author notes:** **Corresponding Author:** Richard J. Schwab, M.D. Division of Sleep Medicine Center for Sleep and Circadian Neurobiology University of Pennsylvania Perelman School of Medicine 3624 Market Street, Suite 205 Philadelphia, PA 19104.

**Keywords:** neck flexion, head flexion, neck extension, head extension, upper airway, soft-tissue movement, obstructive sleep apnea

## Abstract

**Rationale:** Head and neck flexion/extension affect upper airway size. The mechanisms that contribute to these effects are unclear.

**Objectives:** To investigate the changes in airway caliber and movement of the surrounding soft-tissues in apneics and controls during head/neck flexion and extension.

**Methods:** Upper airway MRI was obtained in 24 controls (AHI<5; 1.5±1.5 events/hour) and 33 apneics (AHI≥5; 33.2±28.7 events/hour) with the neck in flexion, extension, and neutral positions. Differences in airway measures and soft-tissue movement were assessed.

**Results:** During extension, controls and apneics showed increased minimum cross-sectional area (CSA) and lateral dimensions in the retropalatal airway (p≤0.007) and increases in all retroglossal airway measures (p≤0.018) compared to neutral position; controls also had increased retropalatal anteroposterior (AP) dimension (p=0.015). During flexion, both groups showed reduced retropalatal lateral dimensions (p≤0.016); controls also had reduced retroglossal CSA (p=0.007). When examining associations with degree of head/neck bending, moving from flexion to extension resulted in increased retropalatal and retroglossal airway sizes (p<0.0001), less lateral wall narrowing (p≤0.002), and more anterosuperior movement of the soft palate and tongue (p≤0.0001). Results were generally consistent in controls and apneics, although each 1° change from flexion to extension resulted in greater increases in retropalatal airway size in controls (interaction p≤0.005).

**Conclusion:** Controls and apneics showed reductions in retropalatal and retroglossal airway caliber during neck flexion and increases during extension, primarily due to movement of the soft palate, tongue, and lateral pharyngeal walls. These data provide important insights into the role of head and neck position on upper airway caliber.

## INTRODUCTION

Obstructive sleep apnea (OSA) is characterized by recurrent closure of the pharyngeal airway during sleep. The pathophysiologic mechanism that causes airway collapse has not yet been fully elucidated. However, studies have demonstrated that changes in upper airway soft tissue structures contribute to the pathogenesis of OSA (1–5). Studies using computed tomography (CT) (2, 5) and magnetic resonance imaging (MRI) (3, 4) have shown that the upper airway of both normal subjects and patients with OSA is smallest in the retropalatal region during sleep. These studies have also demonstrated that airway narrowing in apneics occurs predominantly in the lateral dimension, rather than the anteroposterior dimension. Our group has demonstrated the importance of lateral pharyngeal soft tissue structures in mediating airway caliber during sleep and wakefulness (6), the effects of continuous positive airway pressure (CPAP) (7), and airway changes during respiration (8). Similarly, other researchers have shown that macroglossia (9), increased soft palate length (10), and craniofacial characteristics such as inferior positioning of the hyoid bone and retrognathia (11–14) contribute to OSA risk.

In addition to soft tissue and craniofacial structures, posture has been shown to be an important determinant of upper airway caliber (15) and the frequency and severity of apnea (16–18). It has been widely observed that patients with OSA often have less severe apnea in the lateral decubitus position than the supine position (16–18). A study conducted by Marques et al. demonstrated improvements in pharyngeal patency in the lateral versus the supine sleep position, as quantified by changes in peak inspiratory airflow (19). This finding aligns with older studies on sleep positioning and OSA (20, 21), which suggest that supine positioning results in retropalatal narrowing and subsequent worsening of apneic episodes. Additionally, as reviewed by Omobomi and Quan (22), positional therapy (preventing patients from sleeping in the supine position) is an effective way to manage OSA.

In addition to body posture, neck flexion and extension are thought to change upper airway size. Endoscopic pharyngeal imaging and radiographic observations have shown that cervical extension produces pharyngeal widening, while flexion narrows the airway in both conscious and anesthetized individuals (23, 24). Our study was designed to evaluate the changes in upper airway caliber and geometry that accompany flexion and extension of the neck in normal subjects and patients with OSA. We utilized volumetric MRI and centroid analysis of the pharyngeal soft tissues to quantify changes in retropalatal and retroglossal airway dimensions, as well as movement of the soft palate, lateral walls, and tongue with neck flexion/extension. Based on prior literature (1, 25), we hypothesized that neck extension would result in increased upper airway area, primarily as a result of increased lateral dimensions, and anterior movement of the tongue and soft palate, whereas neck flexion would lead to decreased upper airway area, primarily as a result of decreased lateral dimensions, and posterior movement of the tongue and soft palate. We also hypothesized that the changes in upper airway caliber and movement of the surrounding soft tissues would be greater in apneics than controls.

## METHODS

### Subject Recruitment

Control subjects were recruited using advertisements placed around the University of Pennsylvania campus, and patients with OSA were referred to the study from the Penn Center for Sleep Disorders outpatient practice. Exclusionary criteria for this study included standard MRI contraindications, pregnancy, age under 18 years-old, and weight over 300 pounds (the MRI table limit). All subjects underwent a thorough history and physical examination, including measurements of height and weight. Informed consent outlining the research objectives and potential risks of this study was obtained from all participants. The experimental protocol was approved by the Institutional Review Board of the University of Pennsylvania.

### Polysomnography

Standard polysomnography techniques were performed, as described in previous studies (26). The number of apneas and hypopneas were recorded per hour of sleep to derive the apnea-hypopnea index (AHI). Subjects were considered controls if they had an AHI <5 events/hour and apneics if they had an AHI ≥5 events/hour.

### MR Scanning Protocol

Magnetic resonance imaging (MRI) studies were performed on a 1.5 Tesla scanner (Signa Advantage, Milwaukee, WI) using a receive-only anterior neck coil (General Electric). Images were obtained using conventional spin echo sequences, with the participant’s neck in anatomically neutral, maximally flexed, and maximally extended positions. Neutral head/neck position was defined as the position in which the Frankfurt plane (the plane intersecting the inferior margin of the orbital opening and the superior margin of the acoustic meatus) was perpendicular to the scanning table. Before each scanning sequence, the participant’s head was placed in an appropriate position (either neutral or maximal flexion/extension). Subjects were requested to breathe through their nose with their mouth closed, and to refrain from swallowing during scanning. To avoid inaccuracy in upper airway measurements due to movement or mouth opening, the subject’s head was secured in each position (neutral, flexion, and extension) during the protocol. Data from images with open mouths in any position were excluded from the analysis.

Initially, sagittal localizer scans were performed to confirm an appropriate field-of-view (FOV). Continuous T1 weighted spin echo sagittal sections centered on the upper airway were acquired for each neck position (repetition time [TR] = 600 msec; echo time [TE] = 8 msec; 3 mm slice thickness; number of excitation [NEX] = 1; field of view [FOV] = 20 cm, 256×256 matrix). In addition, continuous T1 weighted spin echo axial sections spanning the top of the hard palate to the larynx were acquired with the same parameter settings.

### Image Processing and Data Analysis

MRI data were analyzed using Amira 5.4.0 software (Amira, VSG, Burlington, MA). The program allowed segmentation of the retropalatal (RP) and retroglossal (RG) regions using spin-echo images. The angle of the hard palate on the midsagittal image relative to the long axis of the MR scanner was measured with each neck maneuver to evaluate the extent of neck extension and flexion (see **Figure 1**). For each neck position, the minimum airway area and anteroposterior (AP) and lateral dimensions at the level of the RP and RG region were measured. AP and lateral airway dimensions were measured on the cross-section containing the minimum area.

**Figure 1:**
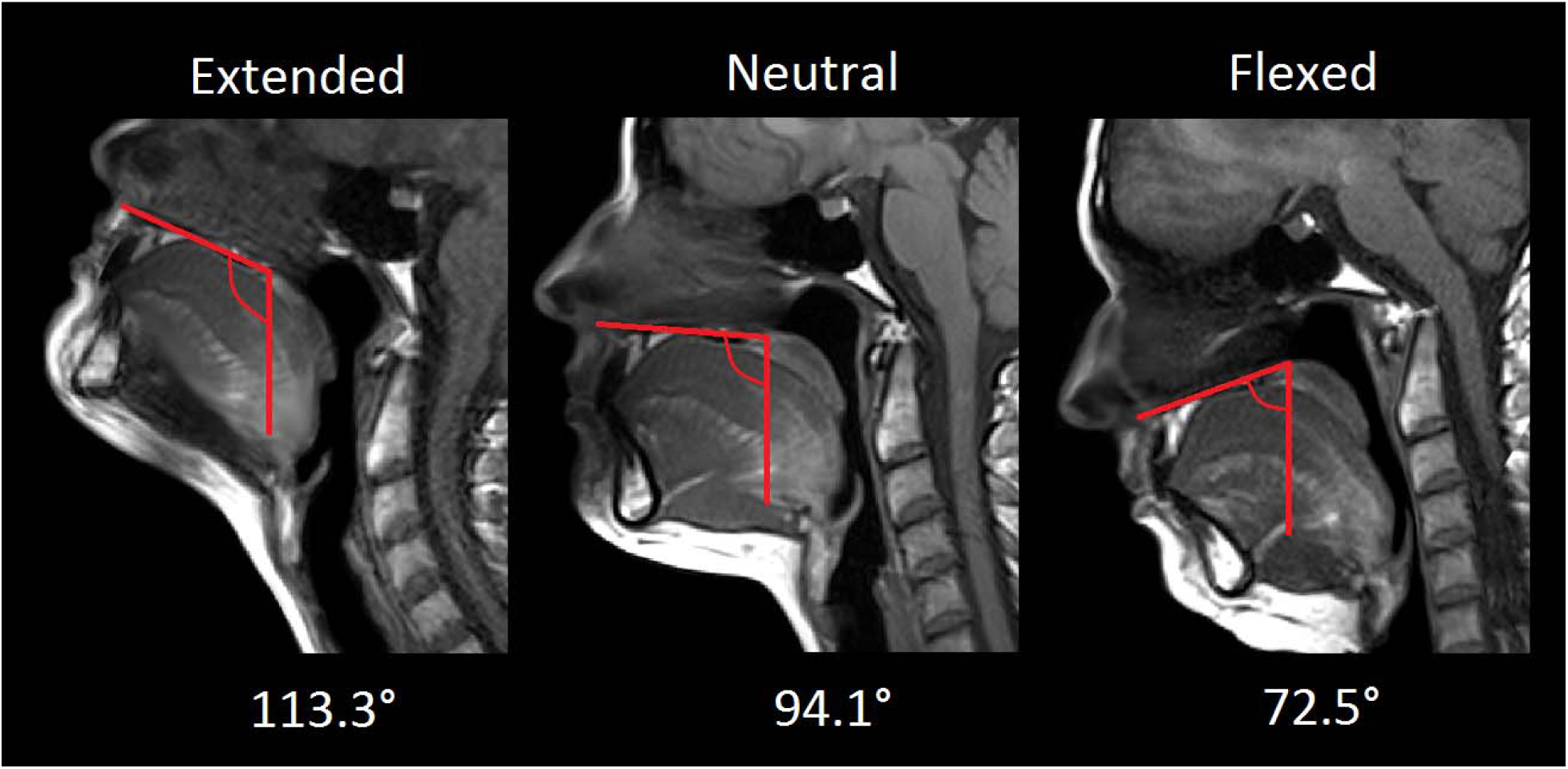
Angle of the hard palate measured with respect to the long axis of the MRI table. Degrees of flexion and extension were calculated relative to the neutral position (middle image). In this example, the patient showed 113.3 – 94.1 = 19.2 degrees of extension (left image) and 94.1 – 72.5 = 21.6 degrees of flexion (right image). In analysis, extension is defined as a positive angle, flexion negative.

On coronal sections, ten surface markers were placed on the left and right sides of the airway wall surface to examine lateral wall movement with neck extension and flexion. These markers included the level of the top of the RP airway, mid-RP airway, RP-RG airway interface, mid-RG airway, and bottom of the RG airway (see **Figure 2**). The airway region of interest was defined from the rostral margin of the soft palate to the base of the tongue.

**Figure 2:**
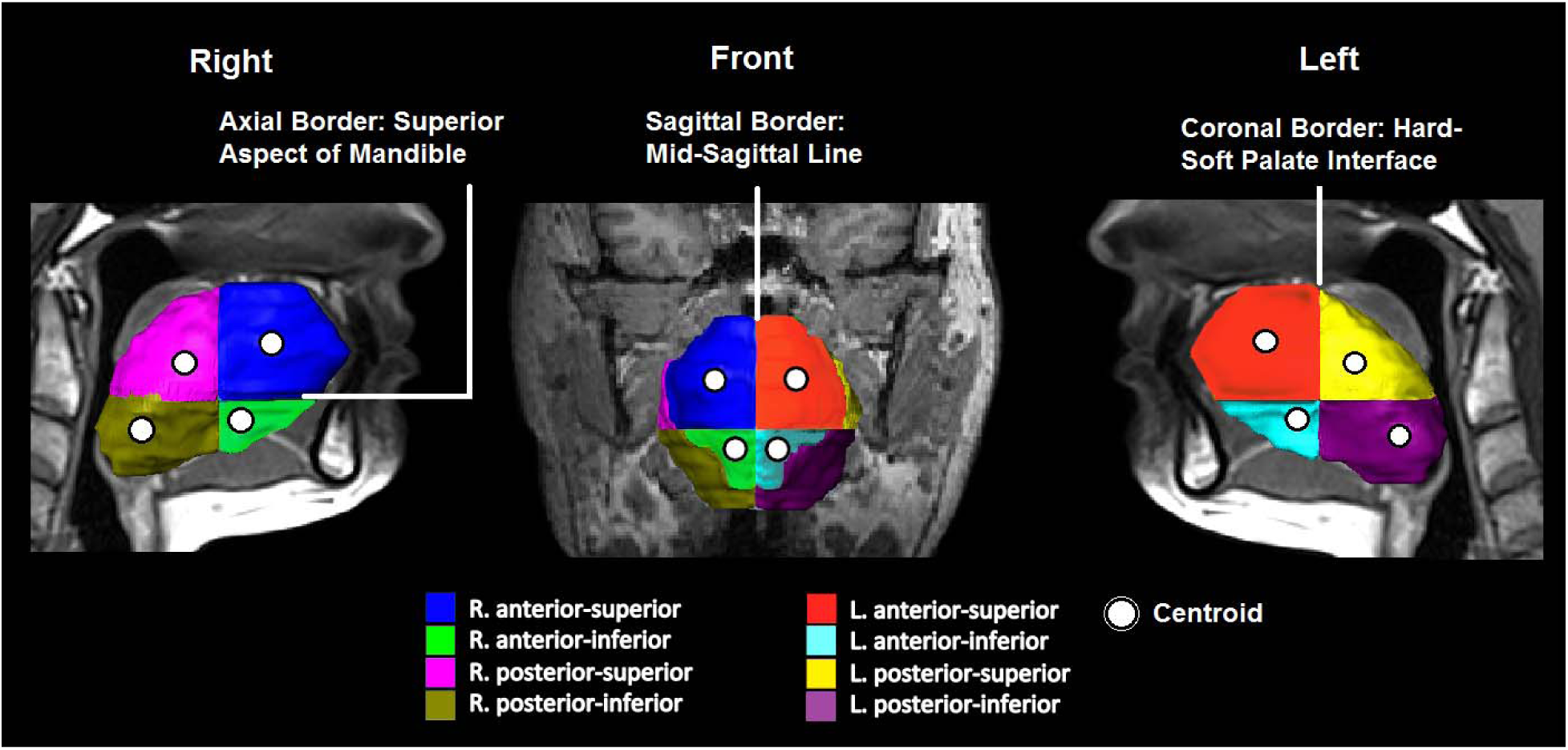
Reconstruction of the tongue and mandible in a control, showing the planes dividing the tongue into 8 octants. White lines represent the planes dividing the tongue into these octants: the axial plane of the superior aspect of the mandibular corpus separates the superior and inferior regions of the tongue; the midsagittal plane separates the left and right regions of the tongue; th coronal plane where the soft and hard palates meet on the posterior nasal spine separates the anterior and posterior tongue. Circles represent the centroid of each tongue octant.

The 3-dimensional centroids of the soft palate and tongue octants (left and right anterior, posterior, superior and inferior regions) were tracked as patients underwent varying degrees of flexion and extension. Tongue centroid position was tracked along 3 axes, with the midsagittal plane separating the left and right regions of the tongue, the coronal plane at the junction of the soft and hard palates (on the posterior nasal spine) separating the anterior and posterior tongue, and the axial plane at the superior aspect of the body of the mandible separating the superior and inferior regions of the tongue (27). Centroid movement due to neck extension or flexion was measured with respect to the MRI table position in each mid-sagittal slice (see **Figure 3**).

**Figure 3:**
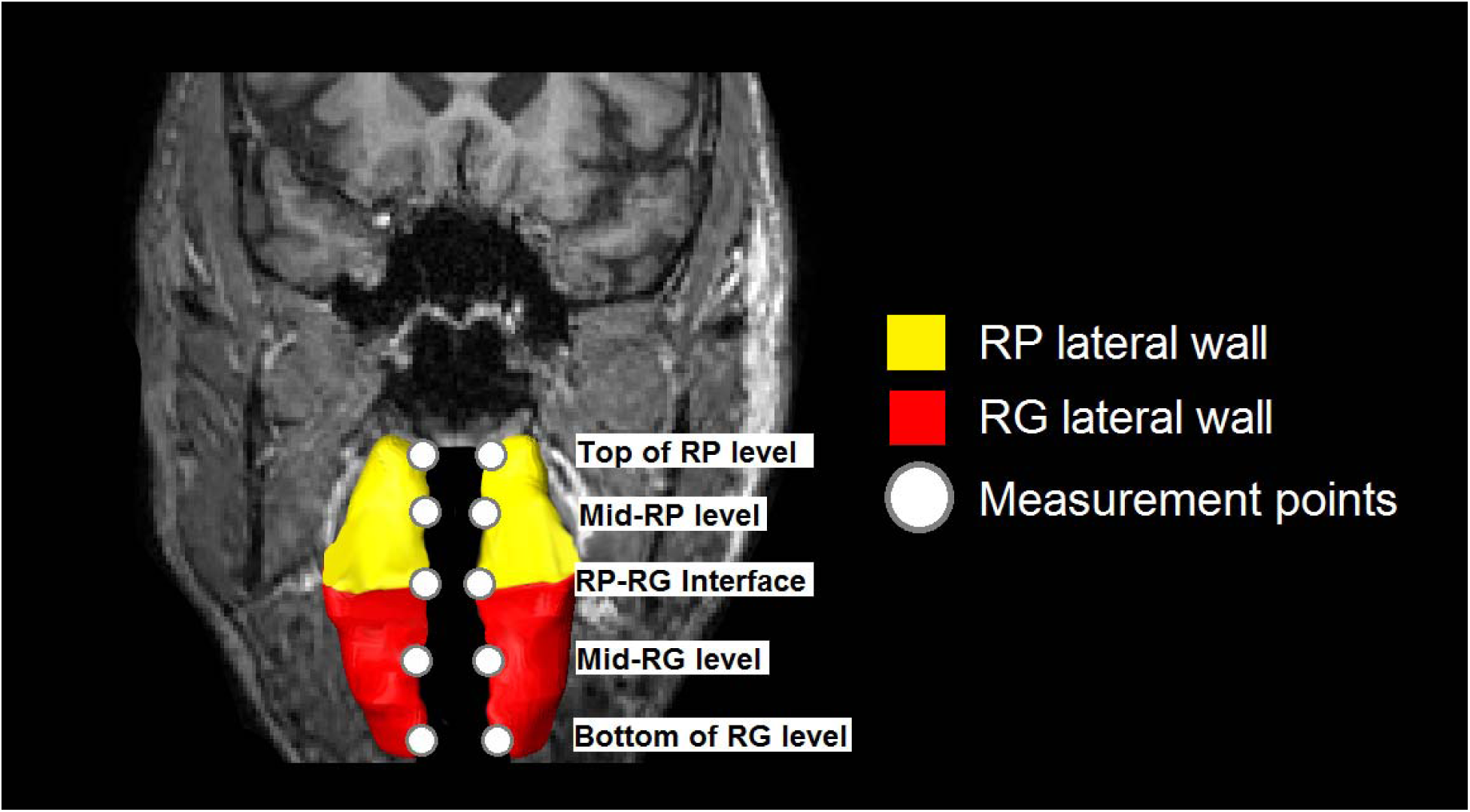
Reconstruction of the retropalatal (RP) and retroglossal (RG) lateral pharyngeal walls with circles indicating the placement of surface markers along the airway wall at five defined points.

### Statistical Methods

Data were summarized using means and standard deviations or frequencies and percentages, unless otherwise specified. Demographic comparisons between apneics and controls were assessed using T-tests (continuous measures) or chi-squared tests (categorical measures). To evaluate whether there were significant changes in airway caliber or soft tissue movement with neck extension/flexion within apneics or controls, we used paired T-tests. Within-subject changes were calculated as values during extension/flexion minus neutral position values. Comparisons of changes in airway caliber or soft tissue movement between apneics and controls were performed using T-tests (unadjusted) or linear regression models adjusted for age, BMI, gender and degree of extension/flexion. To examine associations between the quantitative amount of neck movement and changes in airway caliber or soft tissue movement, we used linear mixed models including measures during neutral, extension, and flexion for each individual; results are presented as the expected change in anatomy for a 1 unit increase in degree of neck extension. Finally, we used statistical interaction tests to evaluate whether the influence of neck extension/flexion on upper airway anatomy differed between apneics and controls. Analyses evaluated the significance of the product term ([degree of neck bending] x [OSA status]) in a linear mixed model that also included the main effects terms.

Statistical significance was determined using the Hochberg step-up procedure (28, 29), applied within specific anatomical domains: *RP airway caliber* (3 measures), *RG airway caliber* (3 measures), *Soft Palate Movement* (2 measures), *Tongue Anteroposterior Movement* (8 measures), *Tongue Superior Movement* (8 measures) and *Lateral Wall Lateral Movement* (5 measures). Any uncorrected p<0.05 was considered nominal evidence of an association. Analyses were performed using Stata/SE 14.2 (StataCorp, College Station, TX).

## RESULTS

### Subject Demographics and Polysomnography Measurements

We performed upper airway imaging during wakefulness in a total of 24 control subjects (AHI<5; mean [±SD] of 1.6±1.5 events/hour) and 33 apneic subjects (AHI≥5; 33.7±29.4 events/hour) in the neutral, extended, and flexed neck positions. As shown in **Table 1**, apneics were significantly older (48.9±11.9 vs. 40.8±14.6 years; p=0.026) and were more obese based on BMI (33.9±4.4 vs. 28.4±5.6 kg/m^2^; p=0.0001) than controls; 57.6% of apneics were males, compared to 37.5% of controls (p=0.134). There were no significant differences in average neck extension (p=0.210) between apneics and controls, although controls showed greater neck flexion (p=0.025).

**Table 1:**
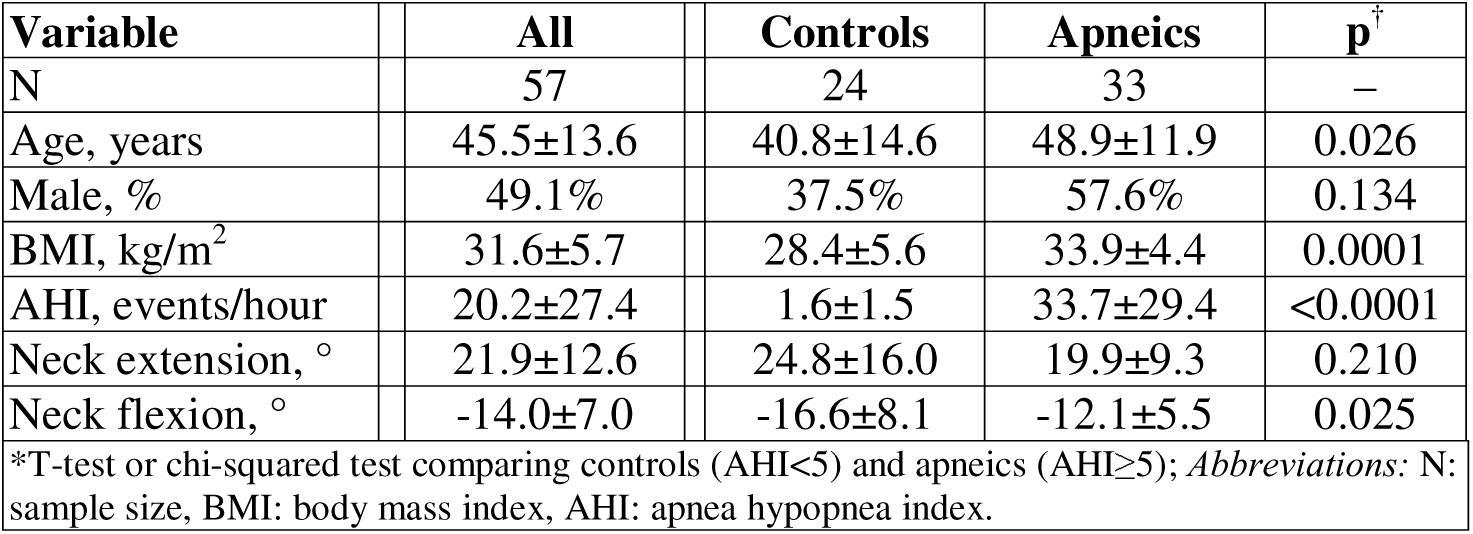
Demographic Summary of Included Apneics and Controls.

| **Variable** | **All** | **Controls** | **Apneics** | **p<sup>†</sup>** |
| --- | --- | --- | --- | --- |
| N | 57 | 24 | 33 | – |
| Age, years | 45.5±13.6 | 40.8±14.6 | 48.9±11.9 | 0.026 |
| Male, % | 49.1% | 37.5% | 57.6% | 0.134 |
| BMI, kg/m<sup>2</sup> | 31.6±5.7 | 28.4±5.6 | 33.9±4.4 | 0.0001 |
| AHI, events/hour | 20.2±27.4 | 1.6±1.5 | 33.7±29.4 | <0.0001 |
| Neck extension, ° | 21.9±12.6 | 24.8±16.0 | 19.9±9.3 | 0.210 |
| Neck flexion, ° | -14.0±7.0 | -16.6±8.1 | -12.1±5.5 | 0.025 |
| *T-test or chi-squared test comparing controls (AHI<5) and apneics (AHI>5); *Abbreviations:* N: sample size, BMI: body mass index, AHI: apnea hypopnea index. |  |  |  |  |

### Regional Airway Changes with Neck Extension and Flexion

We first examined the associations between degree of neck bending (continuous association with degree of neck bending across extension and flexion) and changes in regional airway caliber (**Table 2**); positive increases in degree of neck bending are consistent with the neck becoming less flexed / more extended. Controlling for age, BMI and gender, absolute degree of neck bending was strongly associated with changes in both retropalatal and retroglossal airway measurements among all patients (all p<0.0001). Specifically, within the retropalatal region, we see that each 1° increase in degree of neck bending (e.g., more extension) is expected to increase the minimal cross-sectional area (CSA) by 1.10 mm^2^, anteroposterior (AP) distance by 0.074 mm and lateral distance by 0.132 mm. Within the retroglossal region, each 1° increase in degree of neck bending is expected to increase the minimal cross-sectional area (CSA) by 2.52 mm^2^, AP distance by 0.092 mm and lateral distance by 0.167 mm. The relationship between degree of neck bending and airway changes was different between controls and apneics in the retropalatal region (all interaction p≤0.005), but not the retroglossal region (see **Table 2**). Specifically, increases in the degree of neck bending resulted in larger increases in retropalatal airway measurements among controls than among apneics (see **Figure S1**). Results were consistent in unadjusted analyses (see **Table S1**).

**Table 2:**
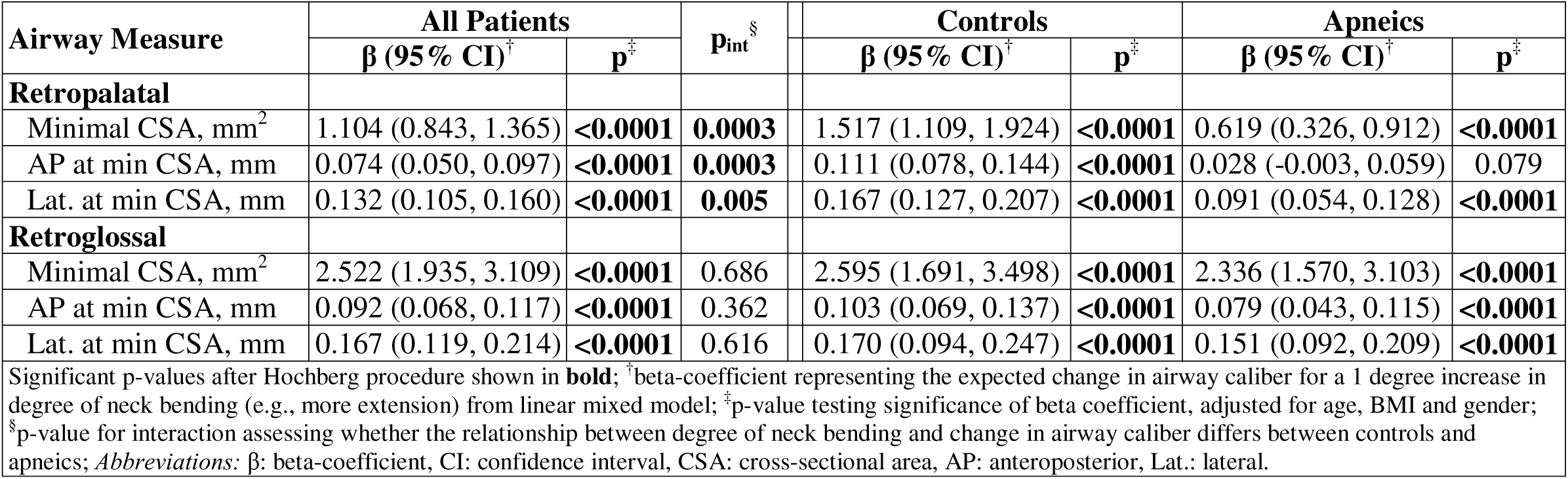
Adjusted Associations between Change in Upper Airway Caliber and Degree of Neck Bending.

In addition to examining the continuous association with degree of neck bending, we examined changes during extension/flexion within and between controls and apneics (see **Table 3**). **Figure 4** shows an increase in both average and minimum cross-sectional area (CSA) in apneics and controls as they transitioned from flexion to neutral to extended neck positions. In the RP region during neck extension, there were significant increases in minimal CSA (p=0.007), AP (p=0.015), and lateral (p=0.005) dimensions among controls and increases in minimal CSA (p=0.002) and lateral dimensions (p=0.001) among apneics. In the RG region during neck extension, both controls and apneics showed significant increases in minimal CSA (p≤0.028), AP (p≤0.004), and lateral (p≤0.017) dimensions. Average magnitude of changes during extension were generally similar between controls and apneics.

**Table 3:** Absolute Change in Upper Airway Measures during Extension or Flexion.

| Airway Measure | Controls |  |  | Apneics |  |  | p<sup>§</sup> | Adj. p<sup>¶</sup> |
| --- | --- | --- | --- | --- | --- | --- | --- | --- |
|  | N | Change<sup>†</sup> | p<sup>‡</sup> | N | Change<sup>†</sup> | p<sup>‡</sup> |  |  |
| Neck Extension |  |  |  |  |  |  |  |  |
| Retropalatal |  |  |  |  |  |  |  |  |
| Minimal CSA, mm<sup>2</sup> | 20 | 38.22±56.35 | **0.007** | 28 | 18.81±28.72 | **0.002** | 0.169 | 0.603 |
| AP at min CSA, mm | 20 | 2.57±4.27 | **0.015** | 28 | 0.93±3.12 | 0.127 | 0.130 | 0.299 |
| Lat. at min CSA, mm | 20 | 3.77±5.25 | **0.005** | 28 | 2.35±3.50 | **0.001** | 0.267 | 0.290 |
| Retroglossal |  |  |  |  |  |  |  |  |
| Minimal CSA, mm<sup>2</sup> | 21 | 68.52±132.54 | **0.028** | 31 | 76.18±78.75 | **<0.0001** | 0.814 | 0.266 |
| AP at min CSA, mm | 21 | 2.96±4.19 | **0.004** | 31 | 3.00±3.48 | **<0.0001** | 0.969 | 0.430 |
| Lat. at min CSA, mm | 21 | 4.83±8.51 | **0.017** | 31 | 4.86±5.59 | **<0.0001** | 0.987 | 0.409 |
| Neck Flexion |  |  |  |  |  |  |  |  |
| Retropalatal |  |  |  |  |  |  |  |  |
| Minimal CSA, mm<sup>2</sup> | 24 | -17.62±42.77 | 0.055 | 31 | -2.84±22.28 | 0.484 | 0.134 | 0.680 |
| AP at min CSA, mm | 24 | -1.27±3.55 | 0.093 | 31 | -0.04±3.12 | 0.951 | 0.175 | 0.749 |
| Lat. at min CSA, mm | 24 | -2.30±3.91 | **0.008** | 31 | -1.14±2.50 | **0.016** | 0.213 | 0.915 |
| Retroglossal |  |  |  |  |  |  |  |  |
| Minimal CSA, mm<sup>2</sup> | 24 | -29.58±48.42 | **0.007** | 29 | -12.49±33.7 | 0.056 | 0.137 | 0.647 |
| AP at min CSA, mm | 24 | -1.19±3.56 | 0.116 | 29 | -0.46±2.85 | 0.389 | 0.414 | 0.945 |
| Lat. at min CSA, mm | 24 | -3.21±6.92 | 0.033 | 29 | -0.84±5.05 | 0.376 | 0.157 | 0.302 |
| Significant p-values after Hochberg procedure shown in **bold**; <sup>†</sup>Mean ± standard deviation change during neck extension or flexion for indicated measure; <sup>‡</sup>p-value from paired T-test assessing whether within group measures during neck extension/flexion are significantly different from neutral position; <sup>§</sup>p-value from T-test comparing whether within group measures during neck extension/flexion are significantly different from between controls and apneics; <sup>¶</sup>p-value comparing changes in controls and apneics, adjusted for age, BMI, gender and degree of neck bending; *Abbreviations*: CSA: cross-sectional area, AP: anteroposterior, Lat.: lateral. |  |  |  |  |  |  |  |  |

**Figure 4:**
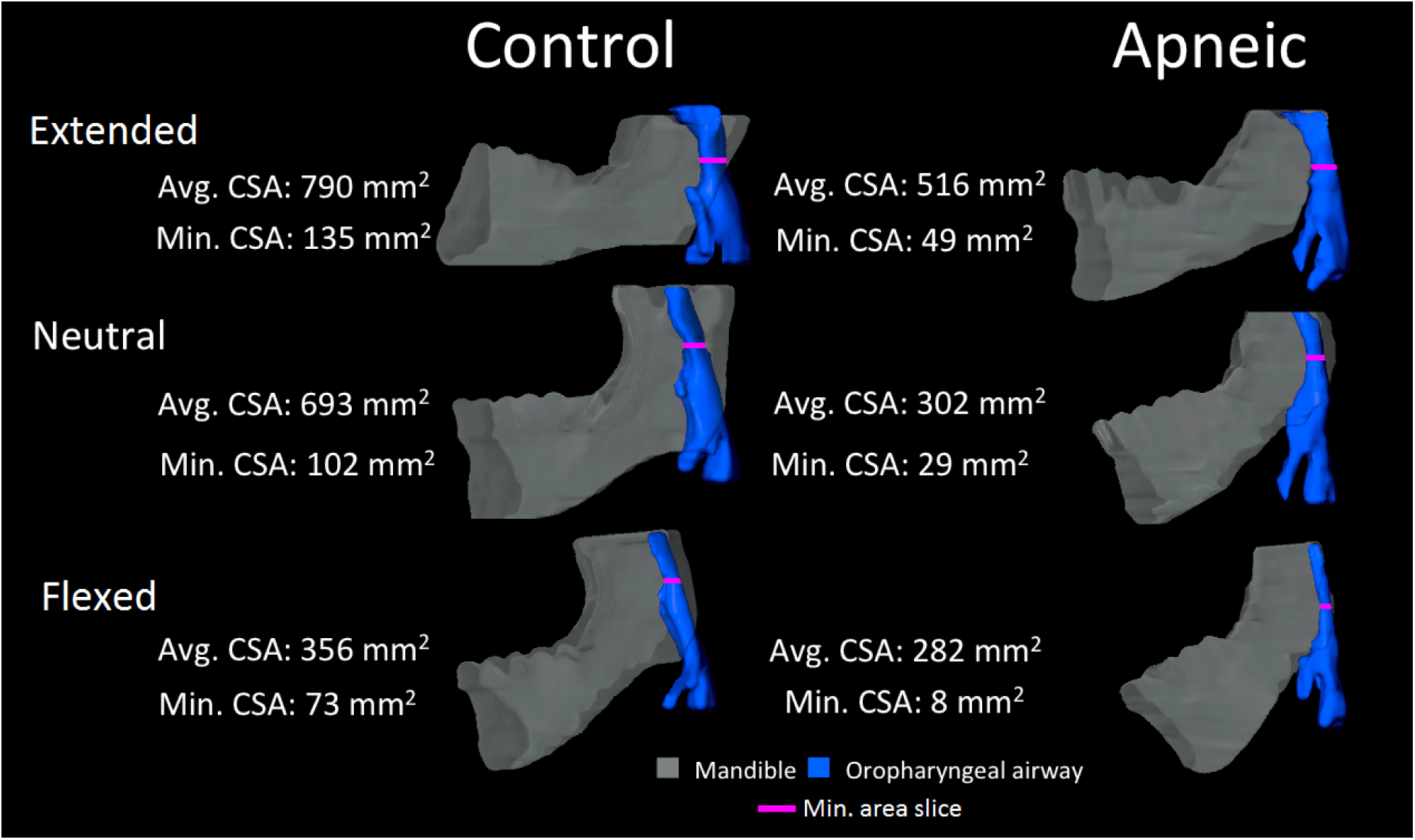
Volumetric reconstruction of the upper airway and mandible during neck extension, neutral position, and flexion in a control and apneic patient. Average and minimum cross-sectional areas are displayed next to each reconstruction, with the points of minimum CSA marked with axial lines.

Similarly, the effect of neck flexion on airway caliber was examined in the RP and RG regions relative to a neutral neck position (see **Table 3** and **Figure 4**). In the RP region during neck flexion, significant decreases in the lateral dimension were seen in both controls (p=0.008) and apneics (p=0.016). In the RG region during neck flexion, controls showed significant decreases in minimal CSA (p=0.007) and nominal decreases in lateral dimension (p=0.033), while apneics showed no statistically significant changes with neck flexion. As with extension, there were no significant differences between controls in apneics in changes during neck flexion.

### Soft Tissue Movement during Neck Extension and Flexion

#### Associations with Degree of Neck Bending

The effect of the continuous degree of neck bending on soft tissue movement was evaluated by examining the centroids of the soft palate, tongue octants and lateral pharyngeal walls. Since extension is defined as a positive angle change, for the tongue and soft palate centroids negative numbers indicate anterior movement, and positive numbers indicate superior movement; for the lateral walls positive numbers indicate lateral movement (airway enlargement) and negative numbers indicate medial movement (airway narrowing). Controlling for age, BMI and gender, we see strong associations between degree of neck extension/flexion and movement in all soft tissue measures among all patients (see **Table 4**); similar results were seen in unadjusted analyses (see **Table S2**). Movement of the soft palate and anterior tongue were similar with increased degree of neck bending, but there was some evidence of less superior tongue movement and less lateral wall movement with more neck bending in apneics compared to controls (multiple interaction p<0.10; see **Table 4**).

**Table 4:**
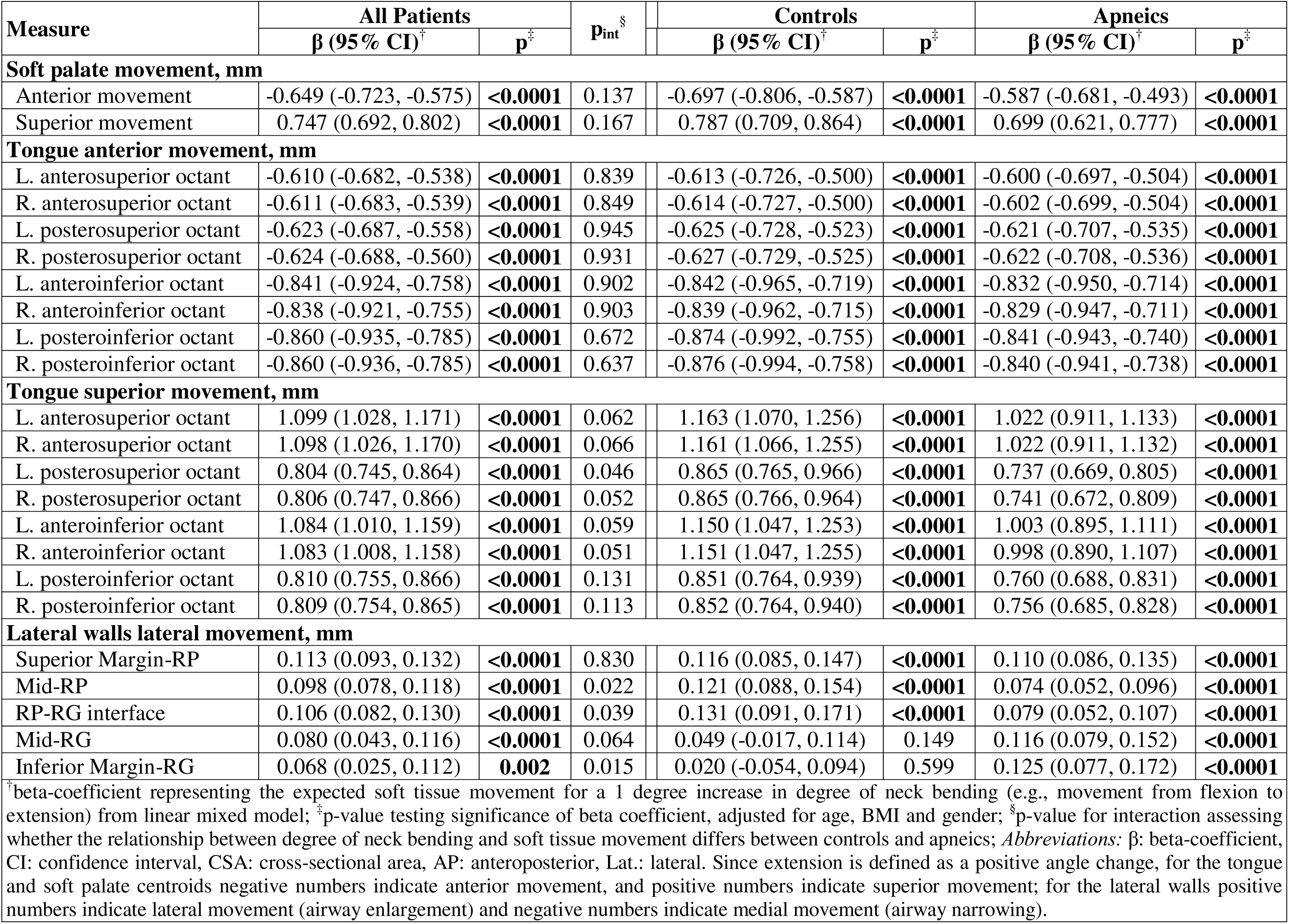
Adjusted Associations between Soft Tissue Movement and Degree of Neck Bending.

More specifically, in covariate adjusted analyses of the soft palate, each 1° increase in neck bending (e.g., moving from flexion to extension) was associated with -0.649 mm more anterior movement (p<0.0001) and 0.747 mm more superior movement (p<0.0001). There was no evidence that these relationships meaningfully differed based on OSA status (all interactions p≥0.137).

When examining the anterior movement of tongue octants, a 1° increase in degree of neck bending (e.g., moving from flexion to extension) resulted in between -0.610 and -0.860 mm more tongue movement (all p<0.0001). There was no significant evidence that this effect differed between apneics and controls (all interactions p≥0.637). When examining the superior movement of tongue octants, for each 1° increase in degree of neck bending, there was between 0.804 and 1.099 mm more superior movement of the tongue. In contrast to the anterior tongue movement, there was some evidence that the superior movement of tongue was generally greater in controls than apneics, with interaction p-values ranging from 0.046 to 0.066 for 6 of the 8 tongue octants (see **Table 4**).

Finally, we examined lateral movement of the lateral pharyngeal walls at five locations: superior margin of the RP airway, mid-RP airway, RP-RG airway interface, mid RG airway, and inferior margin of the RG airway (see **Table 4**). At each location, we observed significant increases in lateral movement associated with increased degree of neck bending (all p<0.002), with estimates ranging from 0.068 to 0.113 mm increased lateral movement for a 1° increase in degree of neck bending. There was again evidence of differences in the effect of neck bending on lateral wall movement between controls and apneics, with more movement in controls than apneics at the mid-RP region (interaction p=0.022) and the RP-RG interface (interaction p=0.039), but less movement in controls than apneics in the Mid-RG region (interaction p=0.064) and inferior margin RG airway (interaction p=0.015).

#### Soft tissue movement in Controls and Apneics during Neck Extension

In addition to associations with quantitative degree of neck bending, we evaluated average changes during neck extension within and between apneics and controls (see **Table 5**). During extension, controls and apneics showed significant superior and anterior movement of the soft palate centroid and the centroids in all the tongue octants (see **Table 5 and Figure 5**). In controls, the soft palate centroid moved superiorly by 15.7 mm (p<0.0001) and anteriorly by 19.7 mm (p<0.0001), while in apneics the soft palate centroid moved superiorly by 12.4 mm (p<0.0001) and anteriorly by 13.2 mm (p<0.0001). In controls, tongue centroid movement during neck extension in all octants ranged from 17.9 to 25.4 mm anteriority (all p<0.0001) and from 17.6 to 26.2 mm superiorly (all p≤0.0001). Similarly, movement in apneics ranged from 13.7 to 20.1 mm anteriorly (all p<0.0001) and from 13.1 to 20.3 mm superiorly (all p<0.0001). There were no significant differences between controls and apneics in either superior or anterior movement of the soft palate or tongue during extension (**Table 5**).

**Table 5:** Soft Tissue Movement during Neck Extension.

| Measure | Controls |  |  | Apneics |  |  | p<sup>§</sup> | Adj. p<sup>¶</sup> |
| --- | --- | --- | --- | --- | --- | --- | --- | --- |
|  | N | Change<sup>†</sup> | p<sup>‡</sup> | N | Change<sup>†</sup> | p<sup>‡</sup> |  |  |
| **Soft palate movement, mm** |  |  |  |  |  |  |  |  |
| Anterior movement | 20 | -19.7±13.01 | < **0.0001** | 27 | -13.24±7.48 | < **0.0001** | 0.057 | 0.507 |
| Superior movement | 20 | 15.73±11.68 | < **0.0001** | 27 | 12.38±7.57 | < **0.0001** | 0.271 | 0.282 |
| **Tongue anterior movement, mm** |  |  |  |  |  |  |  |  |
| L. anterosuperior octant | 19 | -17.88±11.33 | < **0.0001** | 26 | -13.74±7.42 | < **0.0001** | 0.146 | 0.562 |
| R. anterosuperior octant | 19 | -18.13±11.28 | < **0.0001** | 26 | -13.72±7.64 | < **0.0001** | 0.125 | 0.699 |
| L. posterosuperior octant | 20 | -18.39±11.57 | < **0.0001** | 30 | -15±7.05 | < **0.0001** | 0.251 | 0.978 |
| R. posterosuperior octant | 20 | -18.45±11.83 | < **0.0001** | 30 | -15.02±7.16 | < **0.0001** | 0.255 | 0.994 |
| L. anteroinferior octant | 20 | -22.54±14.59 | < **0.0001** | 32 | -19.24±10.19 | < **0.0001** | 0.342 | 0.717 |
| R. anteroinferior octant | 20 | -22.38±14.41 | < **0.0001** | 32 | -19.31±10.13 | < **0.0001** | 0.372 | 0.662 |
| L. posteroinferior octant | 21 | -25.38±14.42 | < **0.0001** | 32 | -20.13±8.86 | < **0.0001** | 0.146 | 0.987 |
| R. posteroinferior octant | 21 | -25.43±14.62 | < **0.0001** | 32 | -20.12±8.83 | < **0.0001** | 0.145 | 0.985 |
| **Tongue superior movement, mm** |  |  |  |  |  |  |  |  |
| L. anterosuperior octant | 19 | 26.2±17.94 | < **0.0001** | 26 | 19.55±10.64 | < **0.0001** | 0.161 | 0.374 |
| R. anterosuperior octant | 19 | 26.2±17.97 | < **0.0001** | 26 | 19.52±10.77 | < **0.0001** | 0.161 | 0.384 |
| L. posterosuperior octant | 20 | 17.66±15.87 | **0.0001** | 30 | 13.12±7.17 | < **0.0001** | 0.242 | 0.408 |
| R. posterosuperior octant | 20 | 17.63±15.95 | **0.0001** | 30 | 13.15±7.16 | < **0.0001** | 0.250 | 0.427 |
| L. anteroinferior octant | 20 | 25.31±18.38 | < **0.0001** | 32 | 20.27±10.68 | < **0.0001** | 0.275 | 0.661 |
| R. anteroinferior octant | 20 | 25.42±18.48 | < **0.0001** | 32 | 20.22±10.63 | < **0.0001** | 0.262 | 0.648 |
| L. posteroinferior octant | 21 | 17.6±12.82 | < **0.0001** | 32 | 14.08±6.87 | < **0.0001** | 0.259 | 0.341 |
| R. posteroinferior octant | 21 | 17.65±12.85 | < **0.0001** | 32 | 14.01±6.71 | < **0.0001** | 0.243 | 0.377 |
| **Lateral walls lateral movement, mm** |  |  |  |  |  |  |  |  |
| Superior Margin-RP | 20 | 2.18±3.96 | 0.024 | 31 | 2.11±2.48 | < **0.0001** | 0.945 | 0.180 |
| Mid-RP | 20 | 2.46±4.47 | 0.024 | 31 | 1.45±2.02 | **0.0004** | 0.353 | 0.293 |
| RP-RG interface | 21 | 2.10±4.68 | 0.053 | 32 | 1.68±2.34 | **0.0003** | 0.701 | 0.212 |
| Mid-RG | 21 | 0.83±10.00 | 0.707 | 32 | 2.53±2.87 | < **0.0001** | 0.457 | 0.039 |
| Inferior Margin-RG | 21 | -0.47±10.30 | 0.836 | 32 | 3.00±3.53 | < **0.0001** | 0.151 | 0.014 |
| Significant p-values after Hochberg procedure shown in **bold**; <sup>†</sup> Mean ± standard deviation change during neck extension for indicated measure; <sup>‡</sup> p-value from paired T-test assessing whether within group change during neck extension is significantly different from zero; <sup>§</sup> p-value from T-test comparing change scores between apneics and controls; <sup>¶</sup> p-value comparing changes in controls and apneics, adjusted for age, BMI, gender and degree of neck extension; *Abbreviations*: L.: left, R.: right, RP: retropalatal, RG: retroglossal. Since extension is defined as a positive angle change, for the tongue and soft palate centroids negative numbers indicate anterior movement, and positive numbers indicate superior movement; for the lateral walls positive numbers indicate lateral movement (airway enlargement) and negative numbers indicate medial movement (airway narrowing). |  |  |  |  |  |  |  |  |

Significant p-values after Hochberg procedure shown in **bold**; <sup>†</sup>Mean ± standard deviation change during neck extension for indicated measure; <sup>‡</sup>p-value from paired T-test assessing whether within group change during neck extension is significantly different from zero; <sup>§</sup>p-value from T-test comparing change scores between apneics and controls; <sup>||</sup>p-value comparing changes in controls and apneics, adjusted for age, BMI, gender and degree of neck extension; *Abbreviations*: L.: left, R.: right, RP: retropalatal, RG: retroglossal. Since extension is defined as a positive angle change, for the tongue and soft palate centroids negative numbers indicate anterior movement, and positive numbers indicate superior movement; for the lateral walls positive numbers indicate lateral movement (airway enlargement) and negative numbers indicate medial movement (airway narrowing).

**Figure 5:**
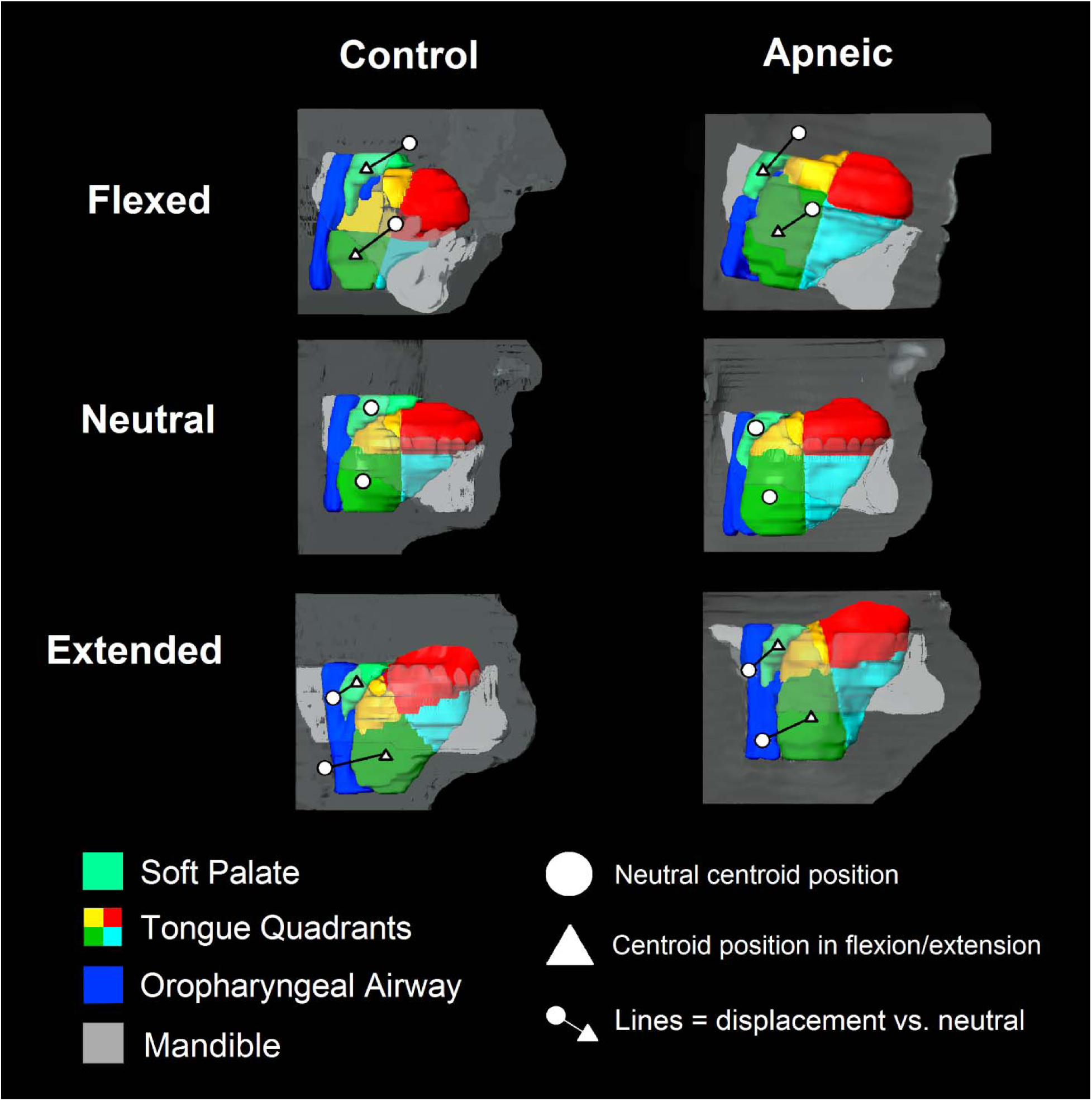
Volumetric reconstruction of the soft palate (teal), right tongue octants (red: anterior-superior, yellow: posterior-superior, light blue: anterior-inferior, and green: posterior-inferior), and upper airway (pink) during flexed, neutral, and extended neck positions in a control and apneic patient. Circles represent centroids of the soft palate and posteroinferior tongue octant in the neutral position. The locations of these points never change in relation to the MRI table, allowing comparison with extended and flexed positions, represented by triangles. Connecting lines demonstrate displacement vs. neutral. During flexion, centroids typically move posteriorly and inferiorly, while during extension they move anteriorly and superiorly. Mandible and profile are included in reference of relative head position.

When examining movement of the lateral walls (**Table 5 and Figure 6**), controls showed nominal evidence of lateral movement from 2.10 to 2.46 mm only in the RP regions (all p≤0.053), while apneics demonstrated significant lateral movement ranging from 1.45 to 3.00 mm during extension across all regions (all p≤0.0004). There was evidence of a difference in the magnitude of movement in lateral walls between apneics and controls in the mid-RG (interaction p=0.039) and the inferior margin RG airway (interaction p=0.014), controlling for covariates and degree of extension.

**Figure 6:**
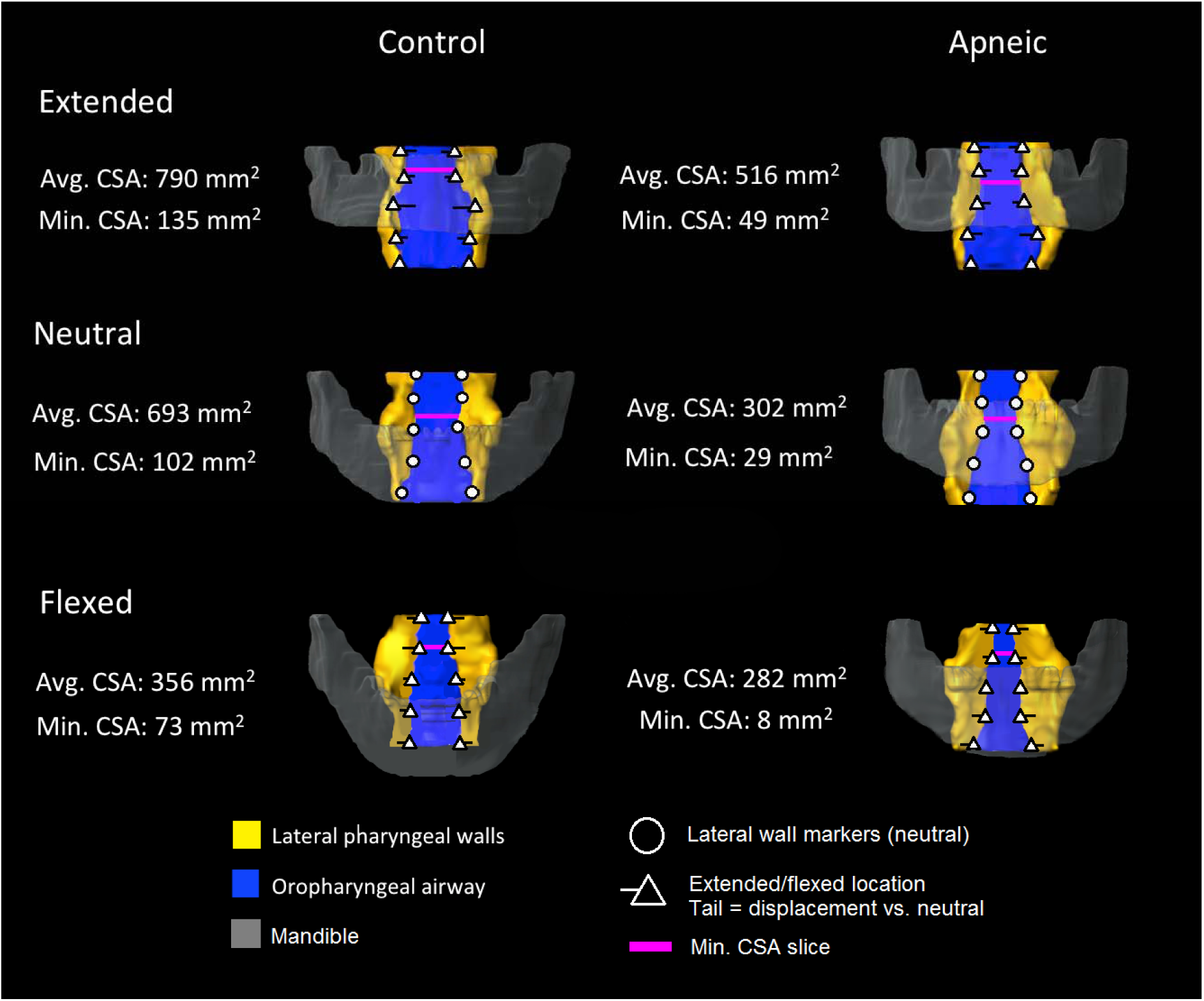
A representative volumetric reconstruction of the upper airway and lateral pharyngeal walls during neck extension (top), neutral position (middle), and flexion (bottom) in a control and apneic patient. Airway caliber is maximized in extension and reduces as the neck flexes, demonstrated by the points located on the lateral walls. Triangular points mark lateral wall position in extension and flexion, with tails indicating the extent of displacement vs. neutral. The points of minimum cross-sectional area are marked with a axial lines.

#### Soft tissue movement in Controls and Apneics during Neck Flexion

When examining soft tissue movement during flexion (see **Table 6 and Figure 6**), as expected, we observed opposite effects of similar magnitude to those seen during neck extension. Both controls and apneics showed significant inferior and posterior movement of the soft palate centroid and the centroid in all tongue octants. In controls, the soft palate centroid moved inferiorly by 17.4 mm (p<0.0001) and posteriorly by 11.8 mm (p=0.0001). Similarly, in apneics, the soft palate centroid moved inferiorly by 12.4 mm (p<0.0001) and posteriorly by 8.0 mm (p<0.0001). Tongue centroid movement in all octants in controls ranged from 9.3 to 16.0 mm posteriorly (all p≤0.0001) and from 16.5 to 22.6 mm inferiorly (all p<0.0001), while centroid movement in apneics ranged from 6.6 to 10.7 mm posteriorly (p<0.0001) and from 12.0 to 16.6 mm inferiorly (all p<0.0001). Similar to extension, there were no significant differences in either inferior or posterior movement of the centroid of the soft palate or tongue during flexion after controlling for covariates between controls and apneics (**Table 6**).

**Table 6:** Soft Tissue Movement during Neck Flexion.

| Measure | Controls |  |  | Apneics |  |  | p<sup>§</sup> | Adj. p<sup>¶</sup> |
| --- | --- | --- | --- | --- | --- | --- | --- | --- |
|  | N | Change<sup>‡</sup> | p<sup>‡</sup> | N | Change<sup>‡</sup> | p<sup>‡</sup> |  |  |
| Soft palate movement, mm |  |  |  |  |  |  |  |  |
| Anterior movement | 23 | 11.79±11.51 | **0.0001** | 31 | 7.96±8.01 | **<0.0001** | 0.155 | 0.714 |
| Superior movement | 23 | -17.35±9.81 | **<0.0001** | 31 | -12.43±5.86 | **<0.0001** | 0.040 | 0.558 |
| Tongue anterior movement, mm |  |  |  |  |  |  |  |  |
| L. anterosuperior octant | 23 | 10.89±10.65 | **0.0001** | 31 | 8.21±7.97 | **<0.0001** | 0.295 | 0.716 |
| R. anterosuperior octant | 23 | 10.70±10.54 | **0.0001** | 31 | 8.22±7.97 | **<0.0001** | 0.330 | 0.759 |
| L. posterosuperior octant | 23 | 9.33±9.24 | **0.0001** | 31 | 6.61±7.75 | **<0.0001** | 0.246 | 0.778 |
| R. posterosuperior octant | 23 | 9.25±9.02 | **0.0001** | 31 | 6.57±7.71 | **<0.0001** | 0.245 | 0.787 |
| L. anteroinferior octant | 23 | 15.81±9.75 | **<0.0001** | 29 | 10.80±9.40 | **<0.0001** | 0.066 | 0.373 |
| R. anteroinferior octant | 23 | 16.01±9.84 | **<0.0001** | 29 | 10.65±9.27 | **<0.0001** | 0.049 | 0.287 |
| L. posteroinferior octant | 23 | 13.23±11.05 | **<0.0001** | 29 | 9.48±8.48 | **<0.0001** | 0.171 | 0.587 |
| R. posteroinferior octant | 23 | 13.24±10.91 | **<0.0001** | 29 | 9.47±8.47 | **<0.0001** | 0.167 | 0.580 |
| Tongue superior movement, mm |  |  |  |  |  |  |  |  |
| L. anterosuperior octant | 23 | -22.17±12.74 | **<0.0001** | 31 | -16.61±8.68 | **<0.0001** | 0.062 | 0.667 |
| R. anterosuperior octant | 23 | -22.12±12.74 | **<0.0001** | 31 | -16.53±8.68 | **<0.0001** | 0.061 | 0.701 |
| L. posterosuperior octant | 23 | -16.58±12.92 | **<0.0001** | 31 | -12.03±7.02 | **<0.0001** | 0.136 | 0.559 |
| R. posterosuperior octant | 23 | -16.51±12.71 | **<0.0001** | 31 | -12.11±7.21 | **<0.0001** | 0.145 | 0.586 |
| L. anteroinferior octant | 23 | -22.48±12.72 | **<0.0001** | 29 | -15.43±8.3 | **<0.0001** | 0.028 | 0.162 |
| R. anteroinferior octant | 23 | -22.58±12.56 | **<0.0001** | 29 | -15.34±8.41 | **<0.0001** | 0.023 | 0.125 |
| L. posteroinferior octant | 23 | -17.44±12.49 | **<0.0001** | 29 | -12.7±6.81 | **<0.0001** | 0.111 | 0.482 |
| R. posteroinferior octant | 23 | -17.45±12.49 | **<0.0001** | 29 | -12.65±6.91 | **<0.0001** | 0.108 | 0.442 |
| Lateral walls lateral movement, mm |  |  |  |  |  |  |  |  |
| Superior Margin-RP | 23 | -2.12±3.88 | **0.016** | 31 | -1.57±2.49 | **0.001** | 0.559 | 0.970 |
| Mid-RP | 23 | -2.22±4.05 | **0.016** | 31 | -1.05±2.02 | **0.007** | 0.215 | 0.915 |
| RP-RG interface | 23 | -3.01±3.83 | **0.001** | 31 | -1.22±2.57 | **0.013** | 0.061 | 0.861 |
| Mid-RG | 23 | -1.61±3.42 | **0.035** | 31 | -1.39±4.03 | 0.064 | 0.837 | 0.980 |
| Inferior Margin-RG | 23 | -3.04±5.45 | **0.014** | 31 | -1.50±4.95 | 0.103 | 0.284 | 0.477 |

Significant p-values after Hochberg procedure shown in **bold**; <sup>‡</sup>Mean ± standard deviation change during neck flexion for indicated measure; <sup>‡</sup>p-value from paired T-test assessing whether within group change during neck flexion is significantly different from zero; <sup>§</sup>p-value from T-test comparing change scores between apneics and controls; <sup>¶</sup>p-value comparing changes in controls and apneics, adjusted for age, BMI, gender and degree of neck flexion; *Abbreviations*: L.: left, R.: right, RP: retropalatal, RG: retroglossal. Since extension is defined as a positive angle change, for the tongue and soft palate centroids negative numbers indicate anterior movement, and positive numbers indicate superior movement; for the lateral walls positive numbers indicate lateral movement (airway enlargement) and negative numbers indicate medial movement (airway narrowing).

During flexion, the lateral airway wall showed medial movement compared to lateral movement in extension. Controls in flexion showed significant medial movement of the lateral walls ranging from 1.61 to 3.04 mm across all regions (all p≤0.035). On the other hand, apneics in flexion showed significant medial movement ranging from 1.05 to 1.57 at the RP levels of the lateral walls, but non-significant movement in the mid-RG and inferior margin RG regions (although the magnitude of movement was comparable to that seen in the RP). Again there were no significant differences in the magnitude of lateral wall movement between controls and apneics (**Table 6**).

## DISCUSSION

We examined changes in upper airway caliber and surrounding soft tissue movement in normal controls and apneics during neck bending maneuvers. Overall, we found that the degree of neck bending significantly influenced upper airway caliber in the retropalatal and retroglossal regions, and resulted in measurable movement of the soft palate, tongue and lateral pharyngeal walls. Neck extension generally resulted in increased airway sizes, superior and anterior movement of the soft palate and tongue octants, and lateral movement at multiple regions of the lateral walls. Conversely, neck flexion lead to reductions in retropalatal and retroglossal airway size, inferior and posterior movement of the soft palate and tongue, and medial movement of the lateral walls. There was some evidence of greater increases in retropalatal airway caliber with neck extension and more superior tongue movement in controls compared to apneics, as well as suggestive differences in lateral wall movement (more movement in mid-RP and RP-RG interface in controls, more movement in mid-RG and inferior margin RG in apneics). Together, these results provide important insights into the significance of head/neck position on upper airway caliber and surrounding soft tissues changes, and the role it may play in OSA pathophysiology. Results highlight the importance of considering neck position when studying the anatomy of the upper airway, determining severity of OSA, and performing overnight sleep studies (e.g., polysomnography).

In addition to evaluating the quantitative effect of increasing the degree of neck bending, we compared average changes from neutral position in apneics and controls during each maneuver (extension or flexion). Both groups showed significant increases in RP and RG airway caliber during neck extension, as well as reductions in the RP lateral dimension and RG minimal CSA measures during neck flexion. In examining anterior soft tissue structure movement, we found that the soft palate and tongue showed anterior and superior movement during extension and posterior and inferior movement during neck flexion in both apneics and controls. Similarly, the lateral pharyngeal walls showed significant lateral movement at the superior margin and mid-RP levels in controls and all levels (RP and RG) in apneics during extension; there was some evidence of more movement in the RG regions in apneics than controls. Conversely, the lateral walls showed significant medial movement at all levels (RP and RG) in controls and all RP but not RG levels in apneics during flexion; there were not significant differences between apneics and controls.

The concept that posture plays a role in determining upper airway size and apnea frequency and severity stems from studies that initially observed that patients with OSA had fewer apneas in the lateral decubitus position than in the supine position (15–18). Using upper airway imaging, Pevernagie and colleagues demonstrated the effects of posture in position-dependent and non-position-dependent awake patients with OSA using electron beam CT (30). Subjects in both groups were found to have similar upper airway dimensions in the prone position. However, in the right side or supine position, the non-position-dependent group showed reduced velopharyngeal cross-sectional area due to reductions in the lateral dimension while the position-dependent group showed increased velopharyngeal CSA due to increases in the lateral dimension. The authors concluded that body position changes in wakefulness principally affect the lateral dimensions of the upper airway.

Although body position plays a significant role in apnea frequency and severity, as evident in positional therapy (17, 18), head/neck position is often overlooked. A study by Zhu and colleagues examined the role of head position independent of trunk position on supine-predominant OSA (31). During overnight polysomnography, the AHI was calculated for total sleep and times spent in various trunk and head positions (supine or lateral). Compared to the complete supine position (trunk supine-head supine), the trunk supine-head lateral position showed a significant reduction in OSA severity. The authors concluded that lateral rotation of the head could reduce AHI even when the trunk remained supine, consistent with similar studies where sleep was natural (32) or drug-induced (33). A study conducted by Tan et. al. went a step further and presented evidence that 30° to 60° head rotations in the supine sleeping position improved airway obstruction in OSA patients compared to no head rotation (34).

Along with head rotation, head/neck flexion and extension are important factors in determining OSA severity. Radiographic studies have shown that cervical extension produces pharyngeal widening, while cervical flexion produces pharyngeal narrowing (23, 24), consistent with our findings. Walsh and colleagues (35) investigated the pharyngeal critical closing pressure (P_crit_) in anesthetized patients during head/neck flexion, extension, and rotation. Relative to the neutral position (where the Frankfurt plane is perpendicular to the bed surface), P_crit_ increased during flexion and decreased during extension. However, P_crit_ was unchanged during head rotation. These findings support our results that head/neck flexion and extension have a marked effect on airway caliber.

The tongue, soft palate, and lateral pharyngeal walls are key upper airway soft tissues that work to maintain pharyngeal airway patency. However, the displacement/movement of these structures through head/neck flexion or extension has been shown to affect upper airway caliber (23). It has been thought that when the chin is pointed downward during flexion, the base of the tongue pushes the base of the epiglottis against the vestibular folds or false vocal cords, potentially leading to narrowing of the larynx (23). Additionally, it has been suggested that a downward displacement of the mandible leads to a reduction of anterior suspensory pull on the hyoid bone, resulting in the hyoid bone dropping inferiorly and posteriorly, reducing the size of the hypopharyngeal airway (36), an effect which may be observed in head/neck flexion. In contrast, extension has been shown to pull the hyoid bone forward due to stretching of the suprahyoid muscles, a behavior which is commonly seen in mouth breathers and could be an important physiological compensation for nasal airway inadequacy (37). Head/neck flexion has also been shown to shorten the pharyngeal airway, which is expected to reduce longitudinal tension in pharyngeal tissue (38). To our knowledge, no studies have examined the length-tension relationship of the upper airway, although studies in elastic tubes and vasculature systems suggest that reduced longitudinal tension leads to increased collapsibility (39, 40). This would support our results that the lateral pharyngeal walls moved to widen the airway during extension and narrow the airway during flexion.

Given that head posture plays a significant role in upper airway patency, it is important to examine it in the context of sleep and polysomnography. The muscles of the soft palate and tongue respond rapidly to negative pressure during wakefulness in order to maintain upper airway patency, although the effectiveness of these reflexes is attenuated by sleep (41). Therefore, it is likely that postural changes such as head/neck flexion could further exacerbate the reduction in airway size observed during sleep. On the other hand, extension could help compensate for this reduction by opening the airway during sleep. Since head/neck position has been shown to influence AHI (31), sleep studies should routinely track head position in addition to body position. Studies have shown significant variability in AHI across nights (42, 43), which could be related to changes in head position. In support of this, a study used a custom-made device that utilizes accelerometry to accurately measure head posture during sleep sessions in the supine position (44). The authors found that when there was an increase in the degree of neck flexion, AHI increased in patients with OSA. Opposite results (decreased neck flexion led to decreased AHI and thus decreased apnea severity) were seen in patients with OSA who were sleeping in the supine position with their heads rotated in either direction. Monitoring head position may be particularly important for patients undergoing a repeat sleep study after intervention for sleep-disordered breathing (e.g., upper airway surgery, an oral appliance, etc.) or in research studies in which AHI is used as an outcome measure (45). Ultimately, data suggest that head/neck position can and should be recorded in all clinical sleep studies.

Contrary to our original hypothesis, we found that the average changes in upper airway anatomy with flexion/extension were generally similar between apneics and controls. We did see that neck movement had a greater impact on the size of the retropalatal airway in controls. as well as suggestive evidence of more superior tongue movement in controls; in both cases, consistent associations were seen in apneics (albeit less strong). Thus, the totality of our evidence suggests that neck flexion will reduce airway caliber and neck extension will increase it in all individuals, regardless of apnea status.

Our study has important strengths and some limitations. Strengths include the detailed measurement of upper airway anatomy size and soft tissue movement, including evaluating the complex movement of the tongue/soft palate with centroids, using standardized procedures in controls and apneics, as well as robust statistical analysis evaluating the proposed hypotheses. While the “true” cross-sectional airway remains parallel to an MRI slice in the neutral position, a potential limitation is that this is not the case in the extended or flexed position. When the head and neck are extended, the airway follows the curve of the spine and is tilted posteriorly, such that the plane of the “true” cross-section rises anteriorly instead of laying horizontal like the MRI slice. Thus each axial slice captures a slightly different cross-section than the “true” cross-section. The same concept applies to head/neck flexion. However, since patients lay supine on an MRI table, changes in airway curvature are limited. Thus, differences between the measured cross-section and “true” cross-section should be minimal. Although there were no differences in degree of extension between apneics and controls, controls showed a significantly greater degree of flexion compared to apneics. This is likely due to anatomical differences between groups, since apneics tend to have a higher BMI and thus had larger submental fat pads. These fat pads likely restricted the range of motion in apneics during neck flexion and serve as an anatomic limitation during neck bending and scanning. To account for these differences when comparing changes between apneics and controls, we adjusted our models for degree of extension/flexion, as well as clinical covariates (age, BMI and gender). Finally, given that the protocol required active participation of the patients (e.g., performing neck extension/flexion), images were necessarily obtained only during wakefulness; while challenging, studies that include careful manipulation of neck/head position during sleep imaging would be useful.

Future studies combining monitoring of head/neck position during polysomnography would provide further insight on the role neck extension and other head postures play on apnea severity. The monitoring of head/neck positions can be achieved through the use of devices similar to the accelerator-based device proposed by Tate et. al (44). Dynamic imaging can be achieved utilizing a recently introduced, rotating MRI table set up proposed by Nguyen et. al

(46). This instrument uses MRI methodology and technology that allows for precise, independent changes in head and body positions to analyze the impact of different angles of head/body rotations on airway caliber.

## CONCLUSIONS

In summary, we have demonstrated significant changes in airway caliber and soft-tissue movement in apneics and controls during neck flexion and extension. Greater degree of neck extension resulted in larger increases in retropalatal and retroglossal airway sizes, more superior and anterior movement of the soft palate and tongue octants, and greater lateral movement of the lateral pharyngeal walls. Conversely, increased neck flexion led to reductions in upper airway caliber, inferior and posterior movement of the soft palate and tongue, and medial lateral wall movement. The effects of degree of flexion/extension on airway caliber or soft tissue movement were generally consistent between apneics and controls. Future studies should examine the effects of neck flexion and extension on polysomnographic traits to elucidate the magnitude of influence these head positions have on disease severity and determine whether or not head position should be included routinely in clinical sleep studies.

## Data Availability

All data produced in the present work are contained in the manuscript

## ONLINE SUPPLEMENT

### SUPPLEMENTAL RESULTS

#### Unadjusted analyses of regional airway changes with neck extension/flexion

Results of analyses examining associations between degree of neck bending and changes in regional airway caliber without covariate adjustment are shown in **Table S1**. Data are very similar to analyses adjusted for age, BMI, and gender (**Table 2**). The degree of neck bending was strongly associated with changes in both retropalatal and retroglossal airway measurements among all patients (all p<0.0001). Within the retropalatal region, we see that each 1° increase in degree of neck bending is expected to increase the minimal-cross sectional area (CSA) by 1.10 mm^2^, anteroposterior (AP) distance by 0.073 mm and lateral distance by 0.132 mm. Within the retroglossal region, each 1° increase in degree of neck bending is expected to increase the minimal cross-sectional area (CSA) by 2.52 mm^2^, AP distance by 0.092 mm and lateral distance by 0.166 mm. When comparing effects between apneics and controls, there were significant interactions in the retropalatal region (p≤0.005), but not the retroglossal region, suggesting differences in the relationship between degree of neck bending and airway caliber in the RP region only.

#### Unadjusted analyses of soft tissue movement with neck extension/flexion

Results of analyses examining associations between degree of neck bending and soft tissue movements (soft palate, tongue octants, and lateral pharyngeal walls) without covariate adjustment are shown in **Table S2**. Data are very similar to primary analyses adjusted for age, BMI, and gender (**Table 4**). Generally, we see strong associations between degree of neck extension/flexion and movement in all soft tissue measures among all patients. Specifically, in covariate adjusted analyses of the soft palate, each 1° increase in degree of neck bending was associated with -0.654 mm more anterior movement (p<0.0001) and 0.748 mm more superior movement (p<0.0001). There was no evidence of effect modification by OSA status on either measure (all p≥0.122), suggesting similar movement in apneics and controls. When examining the anterior movement of tongue octants, a 1° increase in degree of neck bending resulted in between -0.616 and -0.862 mm more movement (all p<0.0001). There was no significant evidence that this effect differed between apneics and controls based on statistical interaction tests (all p≥0.579). Similarly, for each 1° additional increase in neck bending, there was between 0.807 and 1.102 mm more superior movement of the tongue; all measures were greater controls than apneics (with 6 interactions p≤0.05). Finally, at each location on the lateral pharyngeal walls, we observed significant increases in lateral movement associated with increased degree of neck bending (all p<0.002), with estimates ranging from 0.069 to 0.113 mm increased lateral movement for a 1° increase in neck bending. There was evidence of a difference in the relationship between the degree of neck bending on lateral wall movement at the mid-RG region (interaction p=0.020), RP-RG airway interface (interaction p=0.043), and inferior margin of the RG airway (interaction p=0.019) between apneics and controls.

## Author Contributions

Conception and Design: RJS; Analysis and Interpretation: RJS, TCL, ASW, BTK; Drafting of the Manuscript: RJS, TCL, ASW, BTK; Critical revision: RJS, TCL, ASW, SHT, AS, BTK; Final approval of the version to be published: RJS, TCL, ASW, SHT, AS, BTK.

## Subject Code

8.28 Upper Airway: Sleep

## Scientific Knowledge

Posture has been shown to be an important determinant of upper airway caliber, apnea frequency, and apnea severity. Head/neck flexion and extension are also thought to affect upper airway size. Our study quantifies the effects of head/neck flexion and extension on upper airway caliber and surrounding soft-tissue movement using magnetic resonance imaging (MRI) in apneic and control subjects.

## What This Study Adds to this Field

While head/neck flexion and extension have been shown to affect upper airway caliber, the role that the surrounding soft-tissues play is unknown. We have shown that neck extension results in a larger upper airway, due to anterior movement of the soft palate and tongue and lateral movement of the pharyngeal walls. On the other hand, flexion results in a smaller upper airway, due to posterior movement of the soft palate and tongue and medial movement of the pharyngeal walls. Since changes in head position can influence apnea severity, our study supports the implementation of head position tracking during clinical and research sleep studies.

---

**Table S1:** Unadjusted Associations between Change in Upper Airway Caliber and Degree of Neck Bending.

| Airway Measure | All Patients | | $P_{int}^{\S}$ | Controls | | Apneics | |
| --- | --- | --- | --- | --- | --- | --- | --- |
| | $\beta$ (95% CI)<sup>†</sup> | $p^{\ddagger}$ | | $\beta$ (95% CI)<sup>†</sup> | | $\beta$ (95% CI)<sup>†</sup> | $p^{\ddagger}$ |
| **Retropalatal** |  |  |  |  |  |  |  |
| Minimal CSA, mm<sup>2</sup> | 1.104 (0.843, 1.365) | **<0.0001** | **0.0003** | 1.511 (1.104, 1.918) | **<0.0001** | 0.617 (0.324, 0.909) | **<0.0001** |
| AP at min CSA, mm | 0.073 (0.050, 0.097) | **<0.0001** | **0.0003** | 0.110 (0.077, 0.142) | **<0.0001** | 0.028 (-0.003, 0.059) | 0.078 |
| Lat. at min CSA, mm | 0.132 (0.105, 0.160) | **<0.0001** | **0.005** | 0.167 (0.127, 0.207) | **<0.0001** | 0.091 (0.054, 0.127) | **<0.0001** |
| **Retroglossal** |  |  |  |  |  |  |  |
| Minimal CSA, mm<sup>2</sup> | 2.520 (1.933, 3.107) | **<0.0001** | 0.687 | 2.615 (1.719, 3.511) | **<0.0001** | 2.341 (1.575, 3.108) | **<0.0001** |
| AP at min CSA, mm | 0.092 (0.068, 0.117) | **<0.0001** | 0.387 | 0.102 (0.068, 0.136) | **<0.0001** | 0.080 (0.044, 0.116) | **<0.0001** |
| Lat. at min CSA, mm | 0.166 (0.119, 0.213) | **<0.0001** | 0.609 | 0.175 (0.099, 0.251) | **<0.0001** | 0.151 (0.093, 0.210) | **<0.0001** |

Significant p-values after Hochberg procedure shown in **bold**; <sup>†</sup>beta-coefficient representing the expected change in airway caliber measure for a 1 degree increase in degree of neck bending (e.g., more extension) from linear mixed model; <sup>‡</sup>p-value testing significance of beta coefficient; <sup>§</sup>p-value for interaction assessing whether the relationship between degree of neck bending and change in airway caliber differs between controls and apneics; *Abbreviations*: $\beta$ : beta-coefficient, CI: confidence interval, CSA: cross-sectional area, AP: anteroposterior, Lat.: lateral.

**Table S2:** Unadjusted Associations between Soft Tissue Movement and Degree of Neck Bending.

| Measure | All Patients |  | P<sub>int</sub> | Controls |  | Apneics |  |
| --- | --- | --- | --- | --- | --- | --- | --- |
|  | β (95% CI) | p |  | β (95% CI) | p | β (95% CI) | p |
| **Soft palate movement, mm** |  |  |  |  |  |  |  |
| Anterior movement | -0.654 (-0.729, -0.580) | <0.0001 | 0.122 | -0.704 (-0.813, -0.595) | <0.0001 | -0.589 (-0.682, -0.496) | <0.0001 |
| Superior movement | 0.748 (0.694, 0.803) | <0.0001 | 0.154 | 0.788 (0.711, 0.864) | <0.0001 | 0.701 (0.623, 0.778) | <0.0001 |
| **Tongue anterior movement, mm** |  |  |  |  |  |  |  |
| L. anterosuperior octant | -0.616 (-0.688, -0.544) | <0.0001 | 0.735 | -0.627 (-0.739, -0.516) | <0.0001 | -0.602 (-0.697, -0.507) | <0.0001 |
| R. anterosuperior octant | -0.616 (-0.689, -0.544) | <0.0001 | 0.750 | -0.627 (-0.739, -0.516) | <0.0001 | -0.604 (-0.699, -0.508) | <0.0001 |
| L. posterosuperior octant | -0.624 (-0.688, -0.560) | <0.0001 | 0.871 | -0.630 (-0.729, -0.530) | <0.0001 | -0.619 (-0.704, -0.534) | <0.0001 |
| R. posterosuperior octant | -0.626 (-0.690, -0.562) | <0.0001 | 0.853 | -0.632 (-0.731, -0.533) | <0.0001 | -0.620 (-0.705, -0.535) | <0.0001 |
| L. anteroinferior octant | -0.843 (-0.925, -0.761) | <0.0001 | 0.834 | -0.850 (-0.970, -0.731) | <0.0001 | -0.833 (-0.949, -0.716) | <0.0001 |
| R. anteroinferior octant | -0.840 (-0.922, -0.758) | <0.0001 | 0.831 | -0.847 (-0.967, -0.727) | <0.0001 | -0.829 (-0.945, -0.713) | <0.0001 |
| L. posteroinferior octant | -0.862 (-0.936, -0.787) | <0.0001 | 0.614 | -0.880 (-0.995, -0.765) | <0.0001 | -0.841 (-0.941, -0.741) | <0.0001 |
| R. posteroinferior octant | -0.862 (-0.937, -0.787) | <0.0001 | 0.579 | -0.882 (-0.997, -0.767) | <0.0001 | -0.839 (-0.940, -0.739) | <0.0001 |
| **Tongue superior movement, mm** |  |  |  |  |  |  |  |
| L. anterosuperior octant | 1.102 (1.031, 1.174) | <0.0001 | 0.047 | 1.167 (1.077, 1.257) | <0.0001 | 1.022 (0.912, 1.132) | <0.0001 |
| R. anterosuperior octant | 1.101 (1.030, 1.173) | <0.0001 | 0.050 | 1.165 (1.074, 1.256) | <0.0001 | 1.021 (0.911, 1.131) | <0.0001 |
| L. posterosuperior octant | 0.807 (0.748, 0.867) | <0.0001 | 0.031 | 0.869 (0.770, 0.968) | <0.0001 | 0.737 (0.669, 0.805) | <0.0001 |
| R. posterosuperior octant | 0.809 (0.750, 0.868) | <0.0001 | 0.036 | 0.869 (0.772, 0.966) | <0.0001 | 0.741 (0.673, 0.810) | <0.0001 |
| L. anteroinferior octant | 1.085 (1.011, 1.159) | <0.0001 | 0.044 | 1.156 (1.055, 1.257) | <0.0001 | 1.003 (0.896, 1.110) | <0.0001 |
| R. anteroinferior octant | 1.084 (1.009, 1.158) | <0.0001 | 0.037 | 1.158 (1.055, 1.260) | <0.0001 | 0.998 (0.890, 1.105) | <0.0001 |
| L. posteroinferior octant | 0.810 (0.754, 0.865) | <0.0001 | 0.115 | 0.853 (0.766, 0.939) | <0.0001 | 0.760 (0.688, 0.831) | <0.0001 |
| R. posteroinferior octant | 0.809 (0.753, 0.865) | <0.0001 | 0.098 | 0.854 (0.767, 0.940) | <0.0001 | 0.756 (0.685, 0.827) | <0.0001 |
| **Lateral walls lateral movement, mm** |  |  |  |  |  |  |  |
| Superior Margin-RP | 0.113 (0.094, 0.132) | <0.0001 | 0.797 | 0.116 (0.085, 0.147) | <0.0001 | 0.110 (0.085, 0.135) | <0.0001 |
| Mid-RP | 0.099 (0.079, 0.119) | <0.0001 | 0.020 | 0.121 (0.088, 0.154) | <0.0001 | 0.074 (0.052, 0.096) | <0.0001 |
| RP-RG interface | 0.105 (0.082, 0.129) | <0.0001 | 0.043 | 0.129 (0.089, 0.168) | <0.0001 | 0.079 (0.051, 0.106) | <0.0001 |
| Mid-RG | 0.081 (0.044, 0.117) | <0.0001 | 0.079 | 0.051 (-0.014, 0.116) | 0.1266 | 0.115 (0.079, 0.151) | <0.0001 |
| Inferior Margin-RG | 0.069 (0.025, 0.112) | **0.002** | 0.019 | 0.022 (-0.052, 0.096) | 0.5617 | 0.124 (0.076, 0.171) | <0.0001 |

Significant p-values after Hochberg procedure shown in **bold**; <sup>†</sup>beta-coefficient representing the expected soft tissue movement for a 1 degree increase in degree of neck bending (e.g., more extension) from linear mixed model; <sup>‡</sup>p-value testing significance of beta coefficient; <sup>§</sup>p-value for interaction assessing whether the relationship between degree of neck bending and soft tissue movement differs between controls and apneics; *Abbreviations*: β: beta-coefficient, CI: confidence interval, CSA: cross-sectional area, AP: anteroposterior, Lat.: lateral. Since extension is defined as a positive angle change, for the tongue and soft palate centroids negative numbers indicate anterior movement, and positive numbers indicate superior movement; for the lateral walls positive numbers indicate lateral movement (airway enlargement) and negative numbers indicate medial movement (airway narrowing).

**Figure S1.**
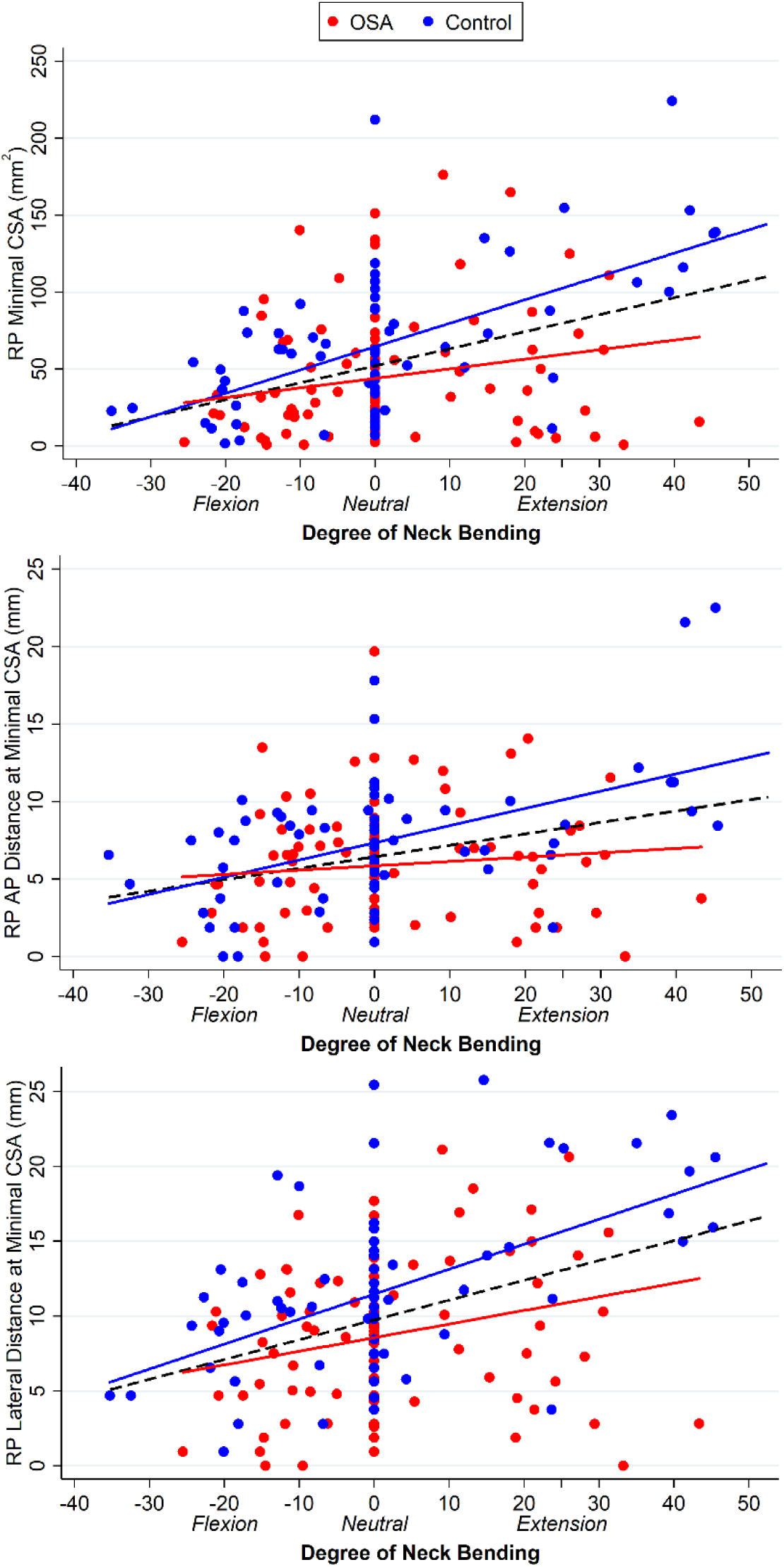
Effect of degree of neck bending on retropalatal airway measurements. Associations between degree of neck bending, where negative values represent flexion relative to neutral and positive values represent extension relative to neutral, are shown overall (black dashed line) and separately in controls (blue line) and apneics (red line). In each retropalatal airway measurement (minimal cross-sectional area [top panel], AP distance [middle panel], and lateral distance [bottom panel]) we observe that increasing the degree of neck bending (e.g., becoming more extended) in controls results in steeper increases in retropalatal airway sizes than among apneics.

**Figure S2.**
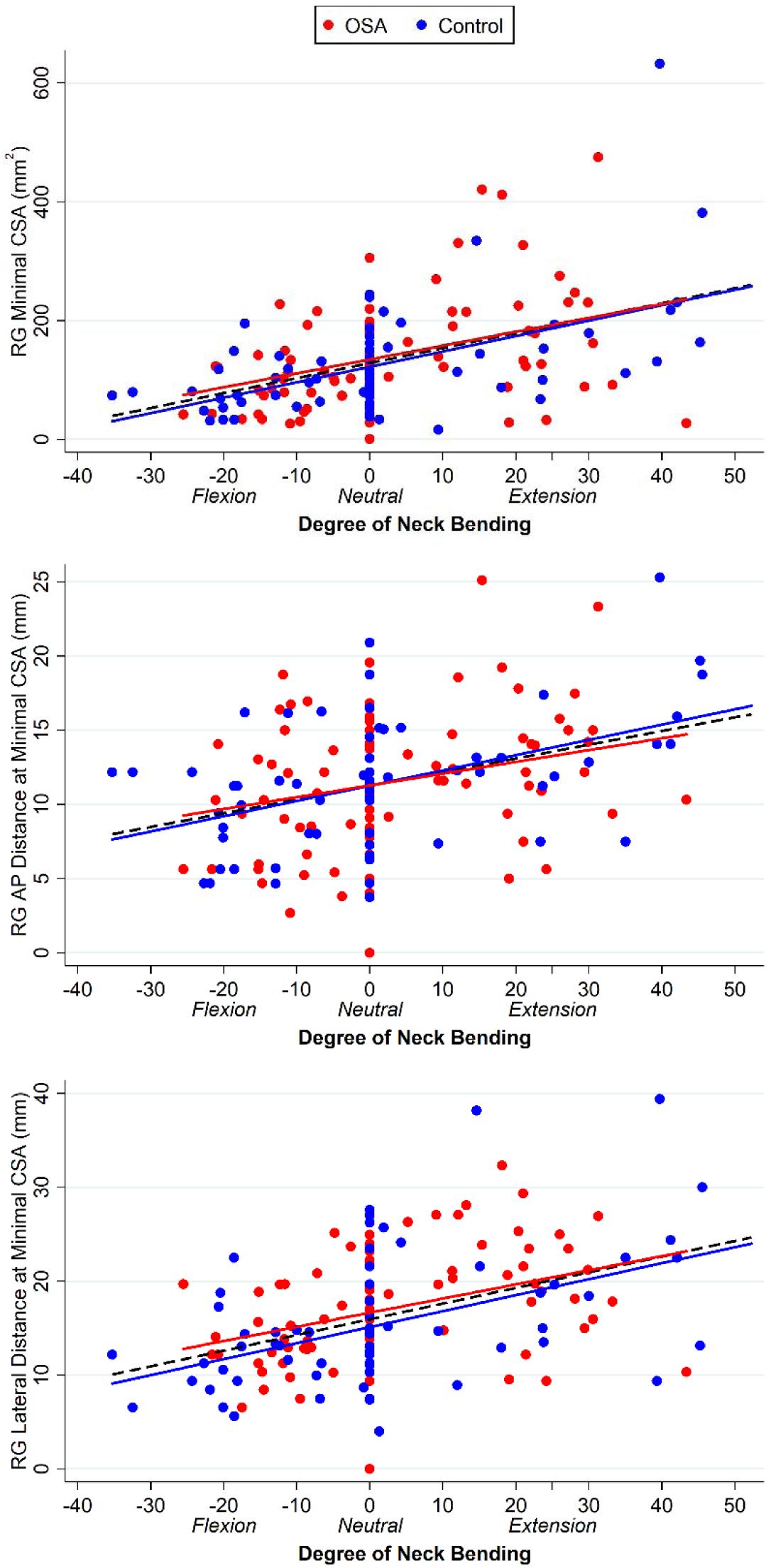
Effect of degree of neck bending on retroglossal airway measurements. Associations between degree of neck bending, where negative values represent flexion relative to neutral and positive values represent extension relative to neutral, are shown overall (black dashed line) and separately in controls (blue line) and apneics (red line). In each retroglossal airway measurement (minimal cross-sectional area [top panel], AP distance [middle panel], and lateral distance [bottom panel]) we observe that increasing the degree of neck bending (e.g., becoming more extended) we see similar changes in retroglossal airway sizes in controls and apneics.

## REFERENCES

1. Schwab RJ, Gupta KB, Gefter WB, Metzger LJ, Hoffman EA, Pack AI. Upper airway and soft tissue anatomy in normal subjects and patients with sleep-disordered breathing. Significance of the lateral pharyngeal walls. Am J Respir Crit Care Med 1995; 152: 1673–1689.

2. Tang XL, Yi HL, Luo HP, Xiong YP, Meng LL, Guan J, Chen B, Yin SK. The application of CT to localize the upper airway obstruction plane in patients with OSAHS. Otolaryngol Head Neck Surg 2012; 147: 1148–1153.

3. Ikeda K, Ogura M, Oshima T, Suzuki H, Higano S, Takahashi S, Kurosawa H, Hida W, Matsuoka H, Takasaka T. Quantitative assessment of the pharyngeal airway by dynamic magnetic resonance imaging in obstructive sleep apnea syndrome. Ann Otol Rhinol Laryngol 2001; 110: 183–189.

4. Faria AC, Garcia LV, Santos AC, Diniz PR, Ribeiro HT, Mello-Filho FV. Comparison of the area of the pharynx during wakefulness and induced sleep in patients with obstructive sleep apnea (OSA). Braz J Otorhinolaryngol 2012; 78: 103–108.

5. Morrell MJ, Arabi Y, Zahn B, Badr MS. Progressive retropalatal narrowing preceding obstructive apnea. Am J Respir Crit Care Med 1998; 158: 1974–1981.

6. Trudo FJ, Gefter WB, Welch KC, Gupta KB, Maislin G, Schwab RJ. State-related changes in upper airway caliber and surrounding soft-tissue structures in normal subjects. Am J Respir Crit Care Med 1998; 158: 1259–1270.

7. Schwab RJ, Pack AI, Gupta KB, Metzger LJ, Oh E, Getsy JE, Hoffman EA, Gefter WB. Upper airway and soft tissue structural changes induced by CPAP in normal subjects. Am J Respir Crit Care Med 1996; 154: 1106–1116.

8. Feng Y, Keenan BT, Wang S, Leinwand S, Wiemken A, Pack AI, Schwab RJ. Dynamic Upper Airway Imaging during Wakefulness in Obese Subjects with and without Sleep Apnea. Am J Respir Crit Care Med 2018; 198: 1435–1443.

9. Barrera JE, Pau CY, Forest VI, Holbrook AB, Popelka GR. Anatomic measures of upper airway structures in obstructive sleep apnea. World J Otorhinolaryngol Head Neck Surg 2017; 3: 85–91.

10. Shigeta Y, Ogawa T, Tomoko I, Clark GT, Enciso R. Soft palate length and upper airway relationship in OSA and non-OSA subjects. Sleep Breath 2010; 14: 353–358.

11. Vieira BB, Itikawa CE, de Almeida LA, Sander HS, Fernandes RM, Anselmo-Lima WT, Valera FC. Cephalometric evaluation of facial pattern and hyoid bone position in children with obstructive sleep apnea syndrome. Int J Pediatr Otorhinolaryngol 2011; 75: 383–386.

12. Gungor AY, Turkkahraman H, Yilmaz HH, Yariktas M. Cephalometric comparison of obstructive sleep apnea patients and healthy controls. Eur J Dent 2013; 7: 48–54.

13. Prabhat KC, Goyal L, Bey A, Maheshwari S. Recent advances in the management of obstructive sleep apnea: The dental perspective. J Nat Sci Biol Med 2012; 3: 113–117.

14. Jo JH, Park JW, Jang JH, Chung JW. Hyoid bone position as an indicator of severe obstructive sleep apnea. BMC pulmonary medicine 2022; 22: 349.

15. Walsh JH, Leigh MS, Paduch A, Maddison KJ, Armstrong JJ, Sampson DD, Hillman DR, Eastwood PR. Effect of body posture on pharyngeal shape and size in adults with and without obstructive sleep apnea. Sleep 2008; 31: 1543–1549.

16. Richard W, Kox D, den Herder C, Laman M, van Tinteren H, de Vries N. The role of sleep position in obstructive sleep apnea syndrome. Eur Arch Otorhinolaryngol 2006; 263: 946–950.

17. Eiseman NA, Westover MB, Ellenbogen JM, Bianchi MT. The impact of body posture and sleep stages on sleep apnea severity in adults. J Clin Sleep Med 2012; 8: 655–666A.

18. Menon A, Kumar M. Influence of body position on severity of obstructive sleep apnea: a systematic review. ISRN Otolaryngol 2013; 2013: 670381.

19. Marques M, Genta PR, Sands SA, Azarbazin A, de Melo C, Taranto-Montemurro L, White DP, Wellman A. Effect of Sleeping Position on Upper Airway Patency in Obstructive Sleep Apnea Is Determined by the Pharyngeal Structure Causing Collapse. Sleep 2017; 40.

20. Cartwright RD, Lloyd S, Lilie J, Kravitz H. Sleep position training as treatment for sleep apnea syndrome: a preliminary study. Sleep 1985; 8: 87–94.

21. Cartwright R, Ristanovic R, Diaz F, Caldarelli D, Alder G. A comparative study of treatments for positional sleep apnea. Sleep 1991; 14: 546–552.

22. Omobomi O, Quan SF. Positional therapy in the management of positional obstructive sleep apnea-a review of the current literature. Sleep Breath 2018; 22: 297–304.

23. Isono S, Tanaka A, Tagaito Y, Ishikawa T, Nishino T. Influences of head positions and bite opening on collapsibility of the passive pharynx. J Appl Physiol (1985) 2004; 97: 339–346.

24. Pirila-Parkkinen K, Pirttiniemi P, Paakko E, Tolonen U, Nieminen P, Lopponen H. Pharyngeal airway in children with sleep-disordered breathing in relation to head posture. Sleep Breath 2012; 16: 737–746.

25. Schwab RJ, Gefter WB, Pack AI, Hoffman EA. Dynamic imaging of the upper airway during respiration in normal subjects. J Appl Physiol 1993; 74: 1504–1514.

26. Schwab RJ, Leinwand SE, Bearn CB, Maislin G, Rao RB, Nagaraja A, Wang S, Keenan BT. Digital Morphometrics: A New Upper Airway Phenotyping Paradigm in OSA. Chest 2017; 152: 330–342.

27. Schwab RJ, Wang SH, Verbraecken J, Vanderveken OM, Van de Heyning P, Vos WG, DeBacker JW, Keenan BT, Ni Q, DeBacker W. Anatomic predictors of response and mechanism of action of upper airway stimulation therapy in patients with obstructive sleep apnea. Sleep 2018; 41.

28. Hochberg Y. A sharper Bonferroni procedure for multiple tests of significance. Biometrika 1988; 75: 800–802.

29. Huang Y, Hsu JC. Hochberg’s Step-up Method: Cutting Corners off Holm’s Step-down Method. Biometrika 2007; 94: 965–975.

30. Pevernagie DA, Stanson AW, Sheedy PF, 2nd, Daniels BK, Shepard JW, Jr. Effects of body position on the upper airway of patients with obstructive sleep apnea. Am J Respir Crit Care Med 1995; 152: 179–185.

31. Zhu K, Bradley TD, Patel M, Alshaer H. Influence of head position on obstructive sleep apnea severity. Sleep Breath 2017; 21: 821–828.

32. van Kesteren ER, van Maanen JP, Hilgevoord AA, Laman DM, de Vries N. Quantitative effects of trunk and head position on the apnea hypopnea index in obstructive sleep apnea. Sleep 2011; 34: 1075–1081.

33. Safiruddin F, Koutsourelakis I, de Vries N. Analysis of the influence of head rotation during drug-induced sleep endoscopy in obstructive sleep apnea. Laryngoscope 2014; 124: 2195–2199.

34. Tan SN, Kim JM, Kim J, Sung CM, Kim HC, Lee J, Lim SC, White DP, Yang HC, Wellman DA. Head rotation improves airway obstruction, especially in patients with less severe obstructive sleep apnea without oropharyngeal collapse. PLoS One 2022; 17: e0268455.

35. Walsh JH, Maddison KJ, Platt PR, Hillman DR, Eastwood PR. Influence of head extension, flexion, and rotation on collapsibility of the passive upper airway. Sleep 2008; 31: 1440–1447.

36. Makofsky HW. Snoring and obstructive sleep apnea: does head posture play a role? Cranio 1997; 15: 68–73.

37. Daly P, Preston CB, Evans WG. Postural response of the head to bite opening in adult males. Am J Orthod 1982; 82: 157–160.

38. Thut DC, Schwartz AR, Roach D, Wise RA, Permutt S, Smith PL. Tracheal and neck position influence upper airway airflow dynamics by altering airway length. J Appl Physiol (1985) 1993; 75: 2084–2090.

39. Elliott EA, Dawson SV. Test of wave-speed theory of flow limitation in elastic tubes. J Appl Physiol Respir Environ Exerc Physiol 1977; 43: 516–522.

40. Sylvester JT, Brower RG, Permutt S. Effects of Hypoxic Vasoconstriction on the Mechanical Interaction between Pulmonary Vessels and Airways. In: Will JA, Dawson CA, Weir EK, Buckner CK, editors. The Pulmonary Circulation in Health and Disease: Academic Press; 1987. p. 321-334.

41. Shorten GD, Armstrong DC, Roy WI, Brown L. Assessment of the effect of head and neck position on upper airway anatomy in sedated paediatric patients using magnetic resonance imaging. Paediatr Anaesth 1995; 5: 243–248.

42. Ahmadi N, Shapiro GK, Chung SA, Shapiro CM. Clinical diagnosis of sleep apnea based on single night of polysomnography vs. two nights of polysomnography. Sleep Breath 2009; 13: 221–226.

43. Levendowski DJ, Zack N, Rao S, Wong K, Gendreau M, Kranzler J, Zavora T, Westbrook PR. Assessment of the test-retest reliability of laboratory polysomnography. Sleep Breath 2009; 13: 163–167.

44. Tate A, Kurup V, Shenoy B, Freakley C, Eastwood PR, Walsh J, Terrill P. Influence of head flexion and rotation on obstructive sleep apnea severity during supine sleep. J Sleep Res 2021; 30: e13286.

45. Schwab RJ. A quantum advance in PSG recordings: the importance of head position in mediating the AHI. Sleep 2011; 34: 985–986.

46. Nguyen HT, Magalang U, Abduljalil A, Elias S, Schmalbrock P, Chandrasekaran P, Rojas S, Emmons K, Ribble D, Knopp MV. MRI-based methodology to monitor the impact of positional changes on the airway caliber in obstructive sleep apnea patients. Magn Reson Imaging 2019; 61: 233–238.

